# Scenario projections of RSV hospitalizations averted due to new immunization programs in King County, Washington, October 2023 to May 2025

**DOI:** 10.1101/2024.12.13.24319008

**Authors:** Chelsea L. Hansen, Lawrence Lee, Samantha J. Bents, Amanda C. Perofsky, Kaiyuan Sun, Lea M. Starita, Amanda Adler, Janet A. Englund, Eric J. Chow, Helen Y. Chu, Cécile Viboud

## Abstract

**Importance:** In 2023 new immunization strategies became available for preventing respiratory syncytial virus (RSV)-associated hospitalizations in infants and older adults. Modeling studies to understand the population-level impact of their use are important for public health planning.

**Objective:** Estimate the hospitalizations averted in 2023-2024 due to new RSV immunizations and provide scenario projections for future seasons.

**Design:** This modeling study used an RSV transmission model calibrated to RSV-diagnosed hospitalizations.

**Setting:** King County, WA, October 2023-May 2025

**Participants:** Population of King County, WA (2.3 million individuals), disaggregated into infant, pediatric, adult, and older adult age groups.

**Exposures:** RSV vaccination for adults aged ≥60 years, maternal RSV vaccination, and long-acting monoclonal antibodies for infants aged <8 months.

**Main Outcomes(s) and Measures(s):** Proportion of RSV-diagnosed hospitalizations averted in adults ≥60 years and infants <1 year.

**Results:** Approximately 25% of older adults and 33% of infants benefited from active or passive immunization during the 2023-2024 RSV season. We estimate that 108 (95% PI 89-154) RSV-diagnosed hospitalizations were averted, with most of the benefit observed in infants <6 months (23% fewer RSV-diagnosed hospitalizations than baseline) and adults ≥75 years (13% fewer RSV-diagnosed hospitalizations). For the 2024-2025 season, optimistic scenarios of high immunization coverage (50% in older adults and 80% in infants) project reductions of 28.9% (95% PI 28.0-29.7) in adults ≥75 years and 61.2% (95% PI 54.2-66.5) in infants <6 months compared to a counterfactual scenario with no immunizations. Targeting infants eligible for catch-up doses of nirsevimab early in the season increased the proportion of RSV-diagnosed hospitalizations averted in infants 6-11 months from 25.7% (95% CI 21.8-29.8) to 38.7% (95%PI 36.2-40.4). If vaccine protection in older adults wanes by 50% in the second year after immunization, the proportion of RSV-diagnosed hospitalizations averted would decrease to 21.1% (95% PI 20.1-22.0) in adults ≥75.

**Conclusions and Relevance:** Our results suggest a modest reduction in RSV-diagnosed hospitalizations during the 2023-2024 season due to limited availability of immunization products, particularly for infants. We project that higher uptake earlier in the season will lead to substantial reductions in RSV hospitalizations in the 2024-2025 season.

**KEY POINTS:** *Question:* How many respiratory syncytial virus (RSV)-diagnosed hospitalizations were averted in King County, WA during the 2023-2024 season due to new active and passive immunizations and how can we optimize disease reduction strategies in future seasons?

*Findings:* We found moderate reductions in RSV hospitalizations during the 2023-2024 season due to modest coverage. With higher levels of coverage earlier in the season more than half of RSV hospitalizations in infants and a quarter of RSV hospitalizations in older adults could be avoided.

*Meaning:* RSV immunizations are a powerful tool for preventing hospitalizations. Modeling studies can support public health strategies to optimize immunization coverage.

## INTRODUCTION

Respiratory syncytial virus (RSV) is a leading cause of pediatric disease burden with 1500-2800 hospitalizations per 100,000 infants <1 year annually in the US.^1–3^ RSV is also responsible for substantial disease burden in older adults, though testing is less frequent and less sensitive in this age group.^1,4^ Prior to 2023 there were no RSV vaccines for adults and the only available prophylaxis against severe RSV disease in infants was palivizumab, which was reserved for very premature newborns and infants with high-risk conditions.^5,6^ In 2023 a new long-acting monoclonal antibody, nirsevimab, was recommended for routine use in infants along with two vaccines for older adults, one of which was also recommended for use in pregnant persons to protect newborns.^7–9^

Observational studies have evaluated the real-world effectiveness of these immunizations over the course of the 2023-2024 season,^10–14^ but modeling studies to understand the population-level impact of their use and guide public health planning remain scarce. Historically there have been many modeling efforts to deliver short-term forecasts and scenario projections for influenza and COVID-19, helping to guide the public health response to these pathogens.^15–17^ In anticipation of new immunization products there has been growing interest in expanding this work to include RSV, ^18–23^ but modeling efforts remain limited, especially at the local scale where decision-making and planning is done. Here we have leveraged routinely collected county public health surveillance data in King County, Washington to develop an RSV transmission model and project the number of RSV-diagnosed hospitalizations averted in King County during the 2023-2024 RSV season due to new RSV immunizations. We provide scenario projections and implications for future RSV seasons.

## METHODS

### RSV Hospitalization and Demographic Data

King County is Washington’s most populous county with approximately 2.3 million individuals and 19 hospitals. More than 80% of all pediatric hospitalizations, including those with diagnosed RSV, occur at Seattle Children’s Hospital (SCH), the region’s largest pediatric hospital. For children <10 years we used hospitalizations at SCH with a positive RSV test (during or preceding admission) from week 26 of 2017 through week 18 of 2024. For individuals ≥10 years we used RSV-diagnosed hospitalizations among King County residents or at King County hospitals from Washington State’s syndromic surveillance platform for the same period (eMethods).^24,25^ Data were aggregated into a weekly all-ages time series and were also summarized by season (defined as week 40 – week 39) and age group (<6 months, 6-11 months, 1-4 years, 5-59 years, 60-74 years, ≥75 years). We rescaled weekly hospitalizations prior to April 2020 to reflect changes in RSV testing and reporting during the COVID-19 pandemic (see eMethods, eFigure 1). We extracted age-specific population sizes, birth rates, and net migration rates for King County, WA using the tidycensus R package.^26^ We used a Washington specific contact matrix described by Mistry et al. to define contacts within and between age classes.^27^

**Figure 1.**
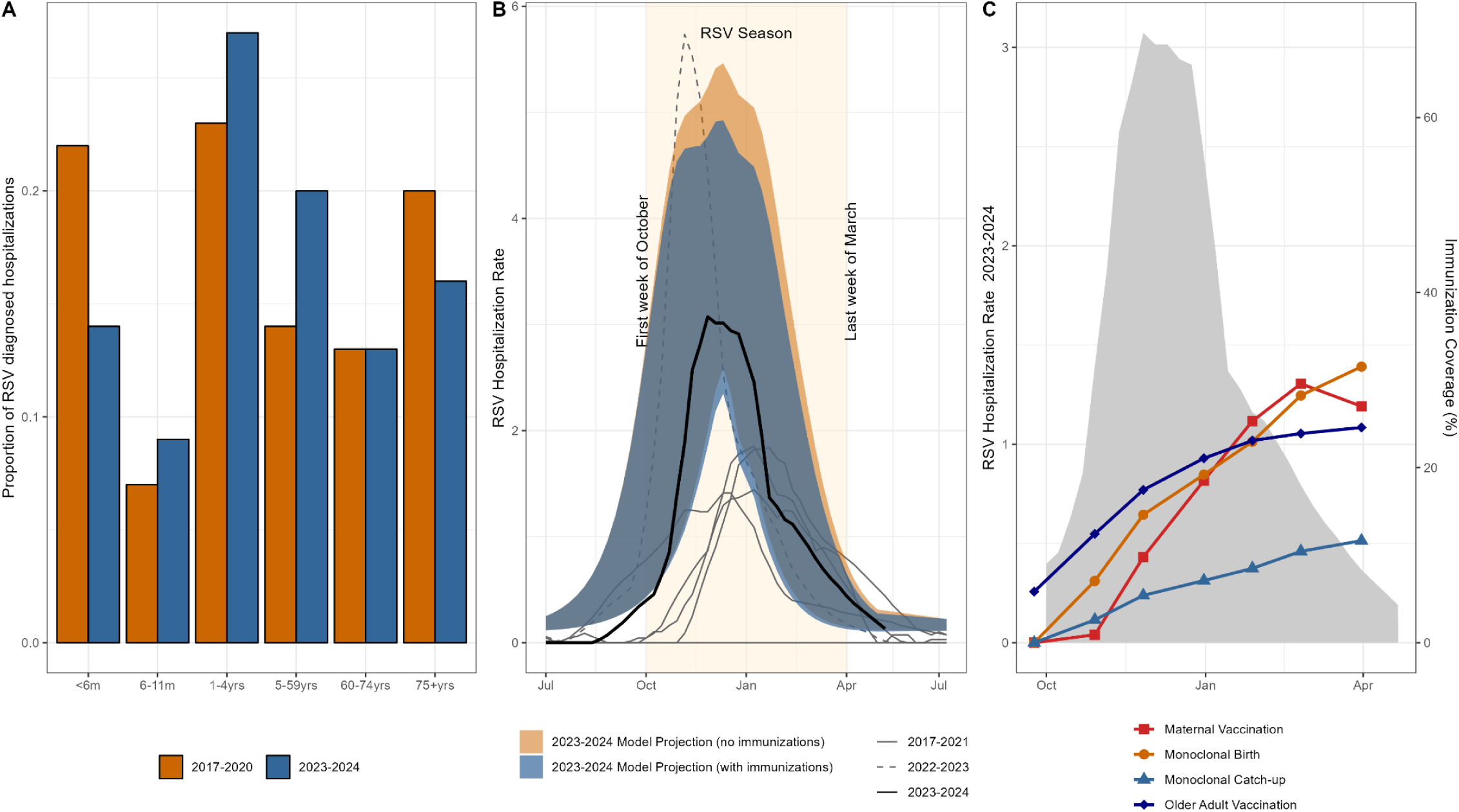
RSV Hospitalization Data from King County, Washington. Panel A) Shows the proportion of RSV-diagnosed hospitalizations in each age group during pre-pandemic seasons (July 2017 to March 2020) (orange) and the 2023-2024 season (October 2023 to April 2024) (blue). Panel B) Shows the weekly RSV hospitalization rate from week 27 (July 1) each year to week 26 (June 30) of the following year from 2017 to 2024. The beige shaded area shows the timing of a typical RSV season: October to March. Prior to the COVID-19 pandemic (2017-2020, solid gray lines) RSV usually peaked in January of each year. The 2022-2023 season had a large, early peak (dashed gray line). The 2023-2024 season peaked in early December (solid black line). The 95% projection intervals (PI) for the model projections are shown in blue (counterfactual) and orange (observed immunization coverage). Panel C) Shows the observed RSV hospitalization rate during the 2023-2024 season in gray. The lines represent the cumulative proportion of the eligible population protected by each strategy: maternal vaccination (red squares), monoclonal antibody catch-up doses (light blue triangles), monoclonal antibody birth doses (orange circles), older adult vaccination (dark blue diamonds).

### Immunization Data

We obtained monthly data on RSV immunizations for King County, WA from July 2023 – April 2024 from the Washington State Immunization Information System (WAIIS).^28^ Data included the doses of nirsevimab for infants 0-7 months and RSV vaccines for individuals 60-74 years and ≥75 years. WAIIS did not have data based on pregnancy status so we assumed that vaccines administered to females between 15 and 49 years were related to pregnancy. We assumed a one-month lag between vaccine administration during pregnancy and infant protection after birth (i.e., doses administered to pregnant persons in September provide protection to infants born in October). We refer to doses of nirsevimab administered to eligible infants born prior to October 1, 2023 as “catch-up doses” and nirsevimab doses administered to infants born during the RSV season as “birth doses.” Additional details on infant immunization coverage are provided in the supplement (eMethods; eTable1).

### RSV Transmission Model and Model Calibration

We developed an age-structured compartmental RSV transmission model adapted from Pitzer et al.^29^ The model assumes short-term maternally-derived partial immunity, repeated infections throughout life, a short period of sterilizing immunity following infection, and a gradual build-up of permanent partial immunity. The model diagram and parameter values are provided in the supplement (eMethods, eTable2, eFigure2, eFigure3). We used maximum likelihood estimation to calibrate the model to the RSV hospitalization data prior to October 1, 2023. Parameters for susceptibility and infectiousness were fixed based on published literature, ^19,21,29^ while parameters for seasonality, reporting rates, and changes in contact patterns during COVID-19 were fitted to King County data (eMethods, eTable 2, eFigure 4).

After fitting the data up to October 1, 2023, we let the model project forward until March 2024, without immunizations. These projections represent our counterfactual scenario (no immunizations) for the 2023-2024 season. We next ran projections for the 2023-2024 season using the reported immunization coverage. We used data from clinical trials and observational studies to set the effectiveness of immunizations against hospitalization (Table 1; eMethods, eTable 2, eFigure 5).^10–14,30–33^ We subtracted these projections from our counterfactual to arrive at an estimated number of RSV-diagnosed hospitalizations averted due to immunizations for the 2023-2024 season.

**Table 1.** Scenarios for the 2024-25 season. Parameters for immunization effectiveness and duration of protection are fixed across scenarios. There are two dimensions of coverage (optimistic and pessimistic) for older adults and infants resulting in four scenarios plus a counterfactual scenario. Coverage levels are based on the observed data in the 2023-24 season with upper bounds based on coverage for other immunizations.

| Scenario Descriptions |  | Vaccination of adults ≥60 years<br>Assuming an average effectiveness of 75% for two years. |  |  |
| --- | --- | --- | --- | --- |
|  |  | <sup>4</sup>**Optimistic Coverage = 50%**<br>≥75 years (67% coverage)<br>60-74 years (43% coverage) | <sup>4</sup>**Pessimistic Coverage = 37.5%**<br>≥75 years (50% coverage)<br>60-74 years (32% coverage) | **Counterfactual**<br>No Coverage in 2024-2025 or 2023-2024 |
| **Long lasting monoclonal antibodies (nirsevimab) for infants <8m**<br>Assuming an average effectiveness of 80% over six months.<br>**Maternal Vaccination**<br>Assuming an average effectiveness of 57% over six months. | <sup>1</sup>**Optimistic Coverage = 80%**<br><sup>2</sup>Catch-up coverage = 74%<br><sup>3</sup>Birth coverage = 87%<br>- Nirsevimab (47%)<br>- Maternal vaccination (40%) | **A** | **B** |  |
|  | **Pessimistic Coverage = 40%**<br><sup>2</sup>Catch-up coverage = 25%<br><sup>3</sup>Birth coverage = 58%<br>- Nirsevimab (31%)<br>- Maternal vaccination (27%) | **C** | **D** |  |
|  | **Counterfactual**<br>No Coverage in the 2024-2025 season. Coverage from the 2023-2024 season does not impact this season. |  |  | **E** |

1. Optimistic coverage for infants (80%) is based on Hepatitis B birth dose coverage in Washington state.<sup>51</sup> 2. There are approximately 13,800 infants born before October 1, 2024 who are eligible for catch-up doses of nirsevimab. Pessimistic coverage for 2024 approximately doubles the observed coverage in the 2023-2024 season. See eTable 1 for more details. 3. There are approximately 11,900 infants born between October 1, 2024 and March 31, 2025. These infants are either born to vaccinated mothers or are eligible for a birth dose of nirsevimab. Pessimistic coverage for the 2024-2025 season is approximately the same as the observed coverage in the 2023-2024 season. See eTable 1 for more details. 4. Coverage includes the observed coverage from the 2023-2024 season (25%). In the optimistic scenario coverage is doubled and in the pessimistic scenario coverage is increased by 50%. Optimistic coverage in the 60-74 year old age group should not exceed 50% (assuming that 50% of this population is eligible due to a high-risk condition<sup>52</sup>). Optimistic coverage for the ≥75 years age group does not exceed influenza vaccine coverage in the ≥65 years age group in Washington state.<sup>53</sup>

### Scenarios for the 2024-2025 Season

For the 2024-2025 season, we established two dimensions of immunization coverage (optimistic and pessimistic) for older adults and infants, resulting in four scenarios plus a counterfactual scenario (Table 1). Following the vaccination guidance for the 2024-2025 season,^34^ we assumed that older adults who were vaccinated in the 2023-2024 season would not get revaccinated in the 2024-2025 season, as vaccine protection is expected to last for at least two years. In the counterfactual scenario we assumed that there were no immunizations for infants or adults ≥60 years in either the 2023-2024 or 2024-2025 seasons. In the pessimistic scenarios we assumed that the cumulative coverage in adults ≥60 years was 37.5% (including the 2023-2024 coverage) and that the combined coverage of nirsevimab and maternal vaccination was 40% in infants. In the optimistic scenarios we assumed 50% coverage in older adults and 80% coverage in infants. See Table 1 for additional details and justifications.

In sensitivity analysis (using the most optimistic scenario – Scenario A), we examined alternative strategies for infant immunization. In scenario A2 the total coverage remained at 80%, but we extended the duration of protection for nirsevimab from 180 days to 270 days. In scenario A3, all catch-up doses of nirsevimab were administered from October to November 2024. In scenario A4 all protection was given via nirsevimab (no maternal vaccination) while in scenario A5, we assumed all newborns were protected by maternal vaccination (no nirsevimab birth doses). In the main analysis for adults ≥60 we assumed that the vaccine remained effective for 2 years and that vaccine protection was against hospitalization given infection but did not prevent infection. In sensitivity analysis we considered vaccine waning, with protection reduced by 50% in the second year after administration. We also considered that vaccination reduced risk of infection.

The study was determined to be non-human subjects research by the University of Washington Institutional Review Board. The study followed the Guidance for the Conduct and Reporting of Modeling and Simulation Studies in the Context of Health Technology Assessment.^35^ Analysis was done in R version 4.3.1.

## RESULTS

### RSV epidemics in King County

The RSV transmission model simulated the population of King County, WA, including 23,700 infants <1 year and 446,500 adults ≥60 years. Despite making up only 20.7% of the total population, these age groups accounted for 62% of the reported RSV hospitalizations in King County from July 1, 2017-April 1, 2020 (Figure 1A). Prior to the COVID-19 pandemic, RSV in King County followed a winter seasonal pattern with peaks typically occurring in January (Figure 1B). Due to public health interventions in response to the COVID-19 pandemic, there was minimal circulation of RSV during the 2020-2021 winter season. As COVID-19 restrictions eased, RSV activity resumed in the summer of 2021, followed by a large peak in November 2022. The timing of the 2023-2024 RSV season was closer to that of pre-pandemic seasons, with a peak in December 2023 (Figure 1B).

### RSV immunizations and hospitalizations during the 2023-2024 winter season

By the end of March 2024, 24.6% of adults ≥60 years had received an RSV vaccine (Figure 1C). For infants born between October 2023 and March 2024, 27.1% were born to vaccinated mothers and an additional 31.1% received a birth dose of nirsevimab, resulting in an overall coverage of 58.2% for newborns. For infants <8 months at the start of the RSV season, 11.4% received a catch-up dose of nirsevimab.^36^ Overall, 33% of all eligible infants were protected by immunization. However, the coverage at the time of the RSV peak in early December was 18.5% for older adults and 21.7% for infants.

In observed hospitalization data for the 2023-2024 RSV season, the RSV-diagnosed hospitalization rate was 38.9 per 100,000 population. This is lower than the observed rate in the 2022-2023 season, but higher than in pre-COVID-19 pandemic seasons (Figure 1B). Our counterfactual model (assuming no immunizations) projected an RSV hospitalization rate of 54.2 (95% projection interval [PI] 47.9 – 69.5), while our immunization model (replicating the observed coverage in this season) projected 49.4 (95% PI 44.0-62.8) RSV hospitalizations per 100,000 population. This amounts to an 8.7% (95% PI 7.4-10.9%) reduction (Table 2), or 108 (95% PI 89-155) RSV-diagnosed hospitalizations averted. The reductions were greater in the age groups targeted by immunizations, including a 13.0% (95% PI 11.4-14.5%) reduction in adults ≥75 years and a 22.9% (95% PI 18.6-27.5%) reduction in infants <6 months (Table 2). There was no difference between scenarios in the 1–59-year age groups, which were not targeted by immunizations. There were 2272 (95% PI 1495 – 2872) vaccinations administered in older adults for each RSV hospitalization averted. In infants, 112 (95% PI 77-147) doses of nirsevimab and 285 (95% PI 147-482) maternal vaccinations were administered for each RSV hospitalization averted.

**Table 2.** Modeled scenario for the 2023-2024 season based on observed immunization coverage and scenario projections for the 2024-25 RSV season (see Table 1).

|  | 2023-2024 Season | 2024-2025 Season |  |  |  |
| --- | --- | --- | --- | --- | --- |
|  | Modeled | Scenario A | Scenario B | Scenario C | Scenario D |
| **RSV hospitalizations averted per 100000** |  |  |  |  |  |
| <6 months | 476.6 (350.7-727.2) | 1362.6 (1029.9-1779.0) | 1362.6 (1029.8-1779.1) | 667.3 (499.5-887.7) | 667.3 (499.5-887.7) |
| 6-11 months | 18.8 (11.7-34.7) | 171.7 (128.5-224.5) | 171.7 (128.4-224.5) | 61.5 (45.8-81.7) | 61.5 (45.8-81.7) |
| 60-74 years | 4.5 (3.5-6.8) | 11.2 (8.6-14.4) | 8.6 (6.6-11.1) | 11.2 (8.6-14.4) | 8.6 (6.6-11.1) |
| ≥75 years | 25.9 (20.0-39.8) | 64.3 (48.4-85.5) | 49.8 (37.4-65.1) | 64.2 (48.4-85.5) | 49.8 (37.4-65.1) |
| All Ages | 4.7 (3.7-7.1) | 13.5 (10.5-17.1) | 12.3 (9.6-15.6) | 9.2 (7.2-11.8) | 8.0 (6.2-10.1) |
| **% of RSV hospitalizations averted** |  |  |  |  |  |
| <6 months | 22.9 (18.6-27.5) | 61.2 (54.2-66.5) | 61.2 (54.2-66.5) | 30.1 (26.1-33.3) | 30.1 (26.1-33.3) |
| 6-11 months | 3.1 (2.0-4.8) | 25.7 (21.8-29.8) | 25.7 (21.8-29.8) | 9.2 (7.7-10.9) | 9.2 (7.7-10.9) |
| 60 - 74 years | 8.4 (7.4-9.4) | 18.8 (18.2-19.4) | 14.4 (14.1-14.8) | 18.8 (18.2-19.4) | 14.4 (14.1-14.8) |
| ≥75 years | 13.0 (11.4-14.5) | 28.9 (28.0-29.7) | 22.2 (21.8-22.6) | 28.9 (28.0-29.7) | 22.2 (21.8-22.6) |
| All Ages | 8.7 (7.4-10.9) | 23.2 (20.7-26.2) | 21.1 (18.7-23.7) | 15.8 (14.2-17.8) | 13.7 (12.4-15.4) |
| **Doses to avert one RSV hospitalization** |  |  |  |  |  |
| Monoclonal antibodies | 112 (77-147) | 105 (93 - 116) |  |  |  |
| Maternal vaccine | 285 (147-482) | 143 (122-165) |  |  |  |
| Older adult vaccination | 2272 (1495-2872) | 1833 (1642-2062) |  |  |  |

### Scenario projections for the 2024-2025 Season

Our most pessimistic scenario for the 2024-2025 season showed reductions in hospitalizations similar to the observed reductions in the 2023-2024 season. Our most optimistic scenario resulted in substantially greater reductions in RSV hospitalizations, including a 61.2% (95% PI 54.2-66.5) reduction in infants <6 months and a 28.9% (95% CI 28.0-29.7) reduction in adults ≥75 years. In our sensitivity analysis testing the impact of different immunization strategies in infants there were minimal differences between strategies for infants <6 months (Figure 2A and 2B). The greatest increase in immunization benefits occurred when all catch-up doses of nirsevimab were administered early in the season, but this mostly benefited infants 6-11 months [from 25.7% (95% CI 21.8-29.8) to 38.7% (95%PI 36.2-40.4) of RSV hospitalizations averted]. In our sensitivity analysis testing waning assumptions in older adults, we found that if vaccine protection is reduced by 50% in the second year the proportion of RSV hospitalizations averted in the optimistic scenario decreases to 21.1% (95% PI 20.1-22.0) among adults >75 years and from 18.8% (95% PI 18.2-19.4) to 13.8% (95% PI 13.1-14.4) among adults 60-74 years (Figure 3A and 3B). If vaccines protect against infection, there is a marginal increase in both the direct benefits to older adults and indirect benefits to other age groups.

**Figure 3.**
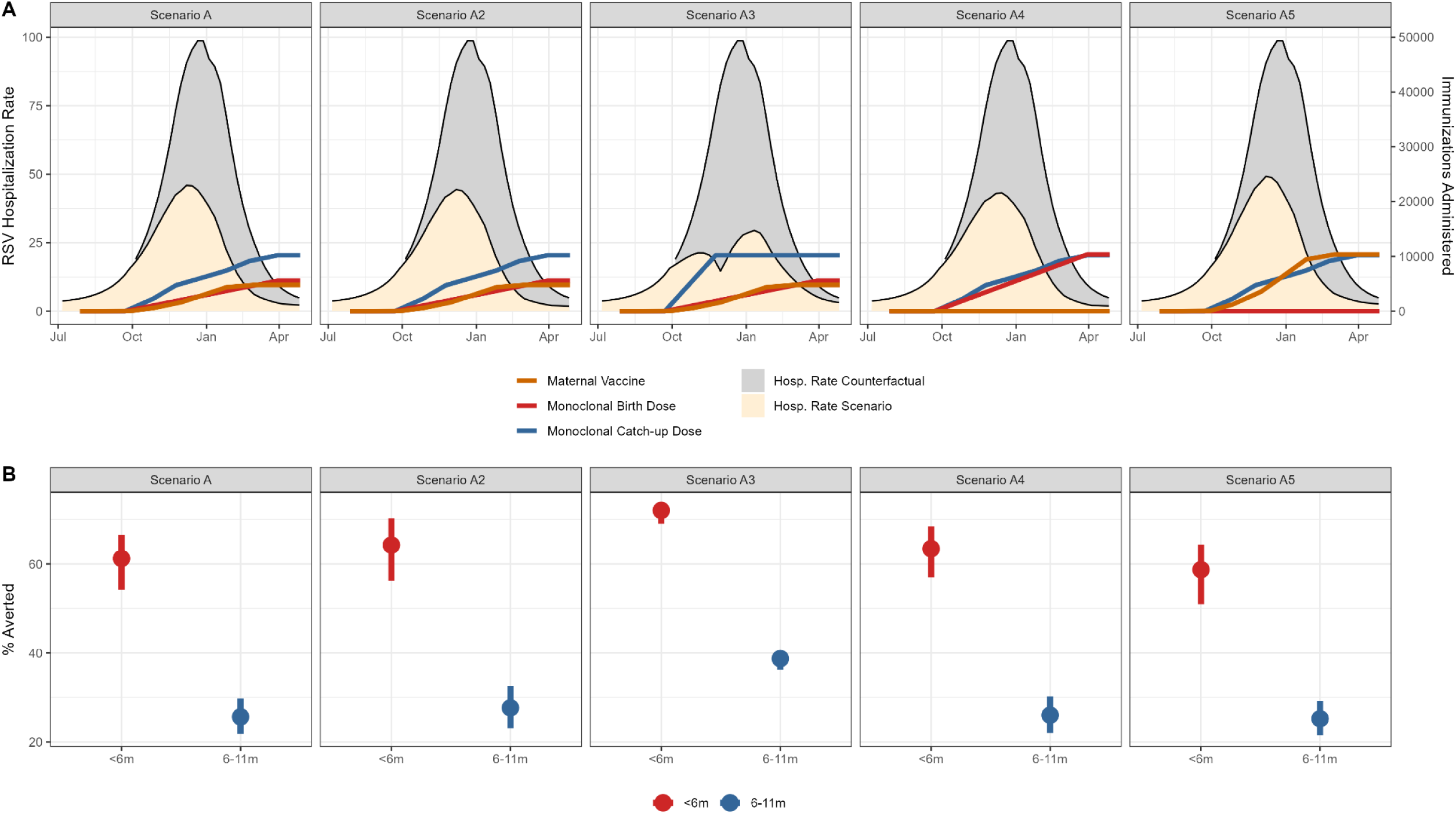
Sensitivity analyses exploring alternative scenarios for infants in the 2024-2025 season. Using the optimistic scenario (Scenario A) for the 2024-25 season we explored alternative immunization strategies in infants, assuming the same total number of infants were immunized (80% of eligible infants). In Scenario A2 all conditions are the same as Scenario A but the duration of protection for monoclonal antibodies is extended from 180 to 270 days. In Scenario A3 the total coverage is the same but all catch-up doses of nirsevimab are administered from October – November. In Scenario A4, all immunization coverage is achieved through nirsevimab (no maternal immunization). In Scenario A5 the catch-up doses of nirsevimab stay the same, but all newborn immunization is achieved through maternal vaccination (no nirsevimab birth doses). In Panel A) the RSV hospitalization rate in infants is shaded gray for the counterfactual scenario of no immunizations and shaded beige for the projected RSV hospitalization rate of each immunization scenario. The cumulative numbers of immunization doses are represented for maternal vaccination (orange), monoclonal birth doses (red) and monoclonal catch-up doses (blue). Panel B) shows the percentage reduction under each immunization scenario for infants <6m and infants 6-11m, compared to the counterfactual scenario of no immunizations.

**Figure 3.**
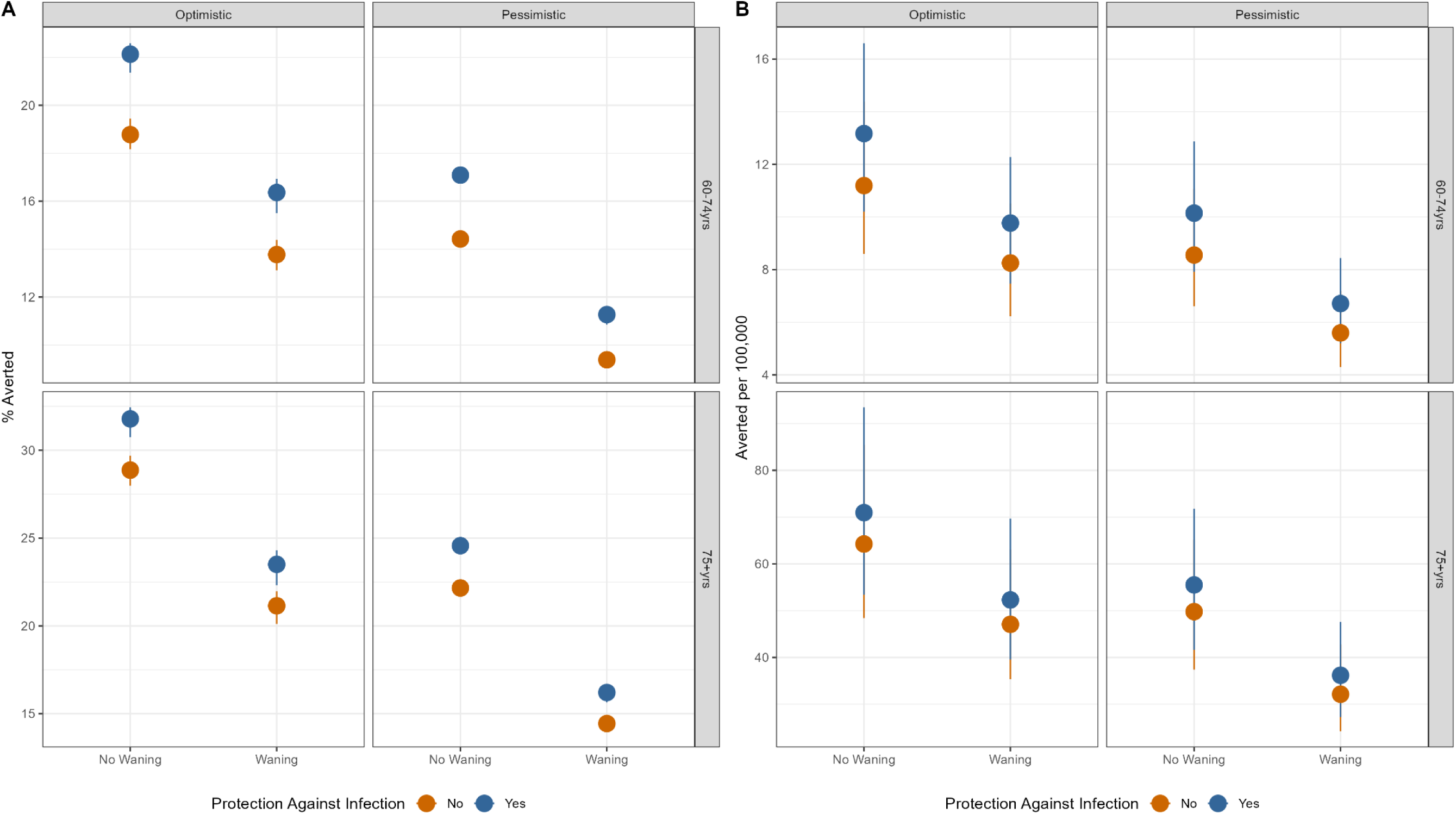
Sensitivity analysis exploring alternative scenarios for older adults in the 2024-2025 season. In our main analysis we assumed that the vaccine protection in older adults was durable for 2 years and that the protection (assuming a VE of 75%) was only against hospitalization given infection and did not protect against infection. In the sensitivity analyses presented here we considered how our estimates would change if protection from the vaccine reduced by 50% in the second year and if the protection was a combination of protection against infection and hospitalization given infection (50% and 50% for a combined relative risk of 0.25, VE=75%). Panel A) Shows the percentage reduction in RSV hospitalizations, compared to the counterfactual scenario of no immunizations, and Panel B) shows the RSV hospitalizations averted per 100,000 population.

## DISCUSSION

We have estimated the number of RSV-diagnosed hospitalizations averted during the 2023-2024 respiratory virus season in King County, WA, due to novel immunization products, and produced scenario projections for the 2024-2025 season. By late March 2024, 25% of eligible older adults and 33% of infants were protected through RSV immunization, although a fraction of these immunizations was administered after peak RSV activity. We found measurable reductions in hospitalizations for the 2023-2024 season, compared to the counterfactual scenario of no immunization: approximately 13% for adults >75 years and 23% for infants <6 months. In our optimistic scenario for the 2024-2025 season, we expect reductions of 30% and 60%, respectively.

During the first years of the COVID-19 pandemic, RSV circulation in King County^37^ and many other locations ^38–43^ was disrupted. Multiple factors likely affected RSV circulation during the pandemic, and as RSV circulation returned, the timing of the RSV season was uncertain. The 2023-2024 RSV season in King County was temporally more similar to seasonal patterns observed prior to the COVID-19 pandemic than during the 2021-2022 or 2022-2023 seasons but was slightly earlier, which likely reduced the impact of immunizations administered late in the season. However, other factors that limited the benefits of immunizations are important to consider, including supply shortages for nirsevimab and confusion around vaccine eligibility for older adults.

Reporting for RSV immunizations is not mandatory, and there is uncertainty in the true level of immunization coverage during the 2023-2024 season, particularly for adults. Publicly available data from the National Immunization Survey (NIS) indicates that 31.9% of adults ≥60 years in WA were vaccinated by the end of March 2024^44^, which is slightly higher than our estimate of 25% for King County. We did not explicitly have data on vaccination among pregnant persons so we assumed doses administered to females 15 to 49 years were because of pregnancy. National data on coverage among pregnant persons indicates that 17.8% of eligible individuals had been vaccinated by the end of January 2024,^45^ which is lower than our estimates for King County, but relies on different methods for ascertaining coverage. We also did not have data on age at the time of nirsevimab administration. Publicly available IIS data on nirsevimab coverage indicates that in WA, 11.4% of infants eligible for catch-up doses of nirsevimab were immunized by March 2024.^36^ We assumed the coverage in King County would be similar to this, and we treated the remaining nirsevimab doses as birth doses, resulting in 31.1% nirsevimab coverage for infants born during the RSV season. This is lower than national data from NIS which indicated 41.3% nirsevimab coverage among infants <8 months in March 2024; however, NIS provides a cross-sectional coverage estimate that may not be directly comparable to the data we used.^46^

In addition to uncertainty regarding the true level of immunization coverage, several other model assumptions may have influenced our results, which we explored through sensitivity analyses. For older adults we explored how the duration of protection and the assumed type of protection – protection against infection versus protection against hospitalization without effect on infection – may change our results. We found that the duration of protection was more important than the type of protection in estimating the number of RSV hospitalizations averted. Likewise, the indirect benefits from reducing transmission in older adults were minimal in the general population. Current data suggests that RSV vaccines for older adults may remain effective for 2-3 years.^47^ Accordingly, at the time of writing it is only recommended that older adults receive a single dose of RSV vaccine.^34,48^ This policy may change as more data become available and modeling studies can help inform recommendations for revaccination in future seasons.

For infants we considered how the duration of protection and timing of nirsevimab administration might impact our results. Assuming that nirsevimab does not provide any protection against infection,^49^ extending the duration of protection did not greatly increase the number of hospitalizations averted. However, this may be a more important consideration in settings where RSV has less pronounced seasonality. We saw the biggest impact in our scenario that administered nirsevimab catch-up doses early in the season, though this was largely confined to reductions in hospitalizations in older infants. For younger infants we considered the tradeoff between maternal vaccination and nirsevimab birth doses. Assuming the same levels of coverage and a similar duration of protection, we found only a marginal difference in favor of nirsevimab (due to higher effectiveness), suggesting that to maximize coverage in the infant population, leveraging all available immunization products is an effective strategy.

### Limitations

Our study has several limitations to consider. First, we calibrated the model to RSV-diagnosed hospitalizations which likely underestimate the true burden of RSV, particularly for adults. However, our model is still useful for understanding the relative differences between scenarios. Additionally, King County is home to many large, highly regarded healthcare facilities which serve patients from an extensive geographic area, and it is likely that some of the RSV-diagnosed hospitalizations in our calibration data are from non-county residents. Separate data from Washington’s syndromic surveillance platform indicate that 15-20% of emergency department visits in King County are from non-residents; a similar proportion may apply to hospitalizations. Although the transmission model simulates the population of King County, we felt that including these non-county residents in the calibration data and resulting scenario projections provided a more complete picture of the RSV burden on the King County hospital system, which has important implications for healthcare capacity planning during the respiratory virus season. Similarly, some of the immunization doses may have been administered to non-county residents.

In May 2024 the CDC updated its recommendations for RSV vaccination in older adults to target all adults ≥75 and only those 60-74 years with high-risk conditions.^34^ While we did look at these age groups separately, we did not model high-risk subpopulations explicitly, nor did we consider the impact of extending vaccines to additional adult populations <60 years. In 2024 a third RSV vaccine option was recommended and made available during the 2024-2025 season.^34,50^ We did not distinguish between products for older adults in our model and going forward, we imagine that any new products would be similar in effectiveness to the ones used this season. And finally, while it is important to extend modeling efforts to the local level, our sample sizes were small and we did not have many historical years of hospitalization data for model calibration, leading to uncertainty in our results. Nevertheless, we were able to fit a transmission model using data sources typically available to local health departments and have provided an example of how partnerships between academic institutions and public health departments can increase modeling capacity at the local level and complement observational studies evaluating vaccine effectiveness.^10^

## CONCLUSIONS

Surveillance data routinely collected by public health agencies can inform transmission models for scenario projections at the local level. Our results suggest a modest reduction in RSV hospitalizations during the 2023-2024 season. Scenario projections for the 2024-2025 season suggest that over 60% of RSV hospitalizations could be averted in infants <6 months with more extensive supplies of immunization products early in the season. Data from observational studies in future seasons will help to validate assumptions regarding the nature and duration of protection and refine scenario projections.

## Disclaimer

The findings and conclusions in this report are those of the authors and do not necessarily represent the official position of the U.S. National Institutes of Health or Department of Health and Human Services.

## Additional contributions

We would like to thank Dr. Drew Bell and Seattle Children’s Hospital and the Washington Department of Health for sharing their data for this analysis. We would like to thank Rebekah Mathew (CSTE), Dr. Catherine Herzog (CDC), and Dr. Matthew Biggerstaff (CDC) for their valuable feedback. We would like to thank Dr. Zhe Zheng (Moderna), Dr. Virginia E. Pitzer (Yale University), and Dr. Daniel M. Weinberger (Yale University) for their earlier work on this topic and for making their code publicly available for other researchers to use and adapt.

## Data availability

The raw data used in this study is not publicly available. Aggregated datasets and model code have been provided on GitHub for reproducibility of results: https://github.com/chelsea-hansen/RSV-Projections-KingCounty.

Model code has been made available as an R package: https://chelsea-hansen.github.io/R.Scenario.Vax/

Researchers wishing to access aggregate data from Seattle Children’s Hospital should contact for information on data sharing agreements. Researcher’s wishing to access Washington’s syndromic surveillance data should contact. Researcher’s wishing to access data from Washington State’s Immunization Information System should contact.

## REFERENCES

1. Zheng Z, Warren JL, Shapiro ED, Pitzer VE, Weinberger DM. Estimated incidence of respiratory hospitalizations attributable to RSV infections across age and socioeconomic groups. Pneumonia (Nathan). 2022;14(1):6.

2. McLaughlin JM, Khan F, Schmitt HJ, et al. Respiratory Syncytial Virus–Associated Hospitalization Rates among US Infants: A Systematic Review and Meta-Analysis. J Infect Dis. 2020;225(6):1100–1111.

3. Rha B, Curns AT, Lively JY, et al. Respiratory Syncytial Virus-Associated Hospitalizations Among Young Children: 2015-2016. Pediatrics. 2020;146(1). doi:10.1542/peds.2019-3611

4. McLaughlin JM, Khan F, Begier E, Swerdlow DL, Jodar L, Falsey AR. Rates of Medically Attended RSV Among US Adults: A Systematic Review and Meta-analysis. Open Forum Infect Dis. 2022;9(7):ofac300.

5. Caserta MT, O’Leary ST, Munoz FM, Ralston SL, COMMITTEE ON INFECTIOUS DISEASES. Palivizumab Prophylaxis in Infants and Young Children at Increased Risk of Hospitalization for Respiratory Syncytial Virus Infection. Pediatrics. 2023;152(1). doi:10.1542/peds.2023-061803

6. Ambrose CS, Chen X, Kumar VR. A population-weighted, condition-adjusted estimate of palivizumab efficacy in preventing RSV-related hospitalizations among US high-risk children. Hum Vaccin Immunother. 2014;10(10):2785–2788.

7. Jones JM, Fleming-Dutra KE, Prill MM, et al. Use of Nirsevimab for the Prevention of Respiratory Syncytial Virus Disease Among Infants and Young Children: Recommendations of the Advisory Committee on Immunization Practices – United States, 2023. MMWR Morb Mortal Wkly Rep. 2023;72(34):920–925.

8. Fleming-Dutra KE, Jones JM, Roper LE, et al. Use of the Pfizer Respiratory Syncytial Virus Vaccine During Pregnancy for the Prevention of Respiratory Syncytial Virus-Associated Lower Respiratory Tract Disease in Infants: Recommendations of the Advisory Committee on Immunization Practices – United States, 2023. MMWR Morb Mortal Wkly Rep. 2023;72(41):1115–1122.

9. Melgar M, Britton A, Roper LE, et al. Use of Respiratory Syncytial Virus Vaccines in Older Adults: Recommendations of the Advisory Committee on Immunization Practices – United States, 2023. MMWR Morb Mortal Wkly Rep. 2023;72(29):793–801.

10. Moline HL, Tannis A, Toepfer AP, et al. Early Estimate of Nirsevimab Effectiveness for Prevention of Respiratory Syncytial Virus-Associated Hospitalization Among Infants Entering Their First Respiratory Syncytial Virus Season – New Vaccine Surveillance Network, October 2023-February 2024. MMWR Morb Mortal Wkly Rep. 2024;73(9):209–214.

11. Paireau J, Durand C, Raimbault S, et al. Nirsevimab Effectiveness Against Cases of Respiratory Syncytial Virus Bronchiolitis Hospitalised in Paediatric Intensive Care Units in France, September 2023-January 2024. Influenza Other Respi Viruses. 2024;18(6):e13311.

12. Ares-Gómez S, Mallah N, Santiago-Pérez MI, et al. Effectiveness and impact of universal prophylaxis with nirsevimab in infants against hospitalisation for respiratory syncytial virus in Galicia, Spain: initial results of a population-based longitudinal study. Lancet Infect Dis. Published online April 30, 2024. doi:10.1016/S1473-3099(24)00215-9

13. Drysdale Simon B., Cathie Katrina, Flamein Florence, et al. Nirsevimab for Prevention of Hospitalizations Due to RSV in Infants. N Engl J Med. 2023;389(26):2425–2435.

14. Assad Zein, Romain Anne-Sophie, Aupiais Camille, et al. Nirsevimab and Hospitalization for RSV Bronchiolitis. N Engl J Med. 2024;391(2):144–154.

15. Borchering RK, Mullany LC, Howerton E, et al. Impact of SARS-CoV-2 vaccination of children ages 5-11 years on COVID-19 disease burden and resilience to new variants in the United States, November 2021-March 2022: a multi-model study. medRxiv. Published online March 10, 2022. doi:10.1101/2022.03.08.22271905

16. Jung SM, Loo SL, Howerton E, et al. Potential impact of annual vaccination with reformulated COVID-19 vaccines: Lessons from the US COVID-19 scenario modeling hub. PLoS Med. 2024;21(4):e1004387.

17. Scenario Modeling Hub. Accessed October 15, 2024. https://scenariomodelinghub.org/index.html

18. RSV Scenario Modeling Hub. Accessed August 16, 2024. https://rsvscenariomodelinghub.org/

19. Hodgson D, Pebody R, Panovska-Griffiths J, Baguelin M, Atkins KE. Evaluating the next generation of RSV intervention strategies: a mathematical modelling study and cost-effectiveness analysis. BMC Med. 2020;18(1):348.

20. Hodgson D, Koltai M, Krauer F, Flasche S, Jit M, Atkins KE. Optimal Respiratory Syncytial Virus intervention programmes using Nirsevimab in England and Wales. Vaccine. 2022;40(49):7151–7157.

21. Zheng Z, Weinberger DM, Pitzer VE. Predicted effectiveness of vaccines and extended half-life monoclonal antibodies against RSV hospitalizations in children. NPJ Vaccines. 2022;7(1):127.

22. Brault A, Pontais I, Enouf V, et al. Effect of nirsevimab on hospitalisations for respiratory syncytial virus bronchiolitis in France, 2023-24: a modelling study. Lancet Child Adolesc Health. 2024;8(10):721–729.

23. van Boven M, Teirlinck AC, Meijer A, et al. Estimating transmission parameters for respiratory syncytial virus and predicting the impact of maternal and pediatric vaccination. J Infect Dis. 2020;222(Suppl 7):S688–S694.

24. Washington State Department of Health. Syndromic Surveillance (RHINO). Accessed September 16, 2024. https://doh.wa.gov/public-health-provider-resources/healthcare-professions-and-facilities/data-exchange/syndromic-surveillance-rhino

25. CDC. Companion Guide: NSSP ED Data on Respiratory Illness. Accessed October 4, 2024. https://www.cdc.gov/nssp/php/onboarding-resources/companion-guide-ed-data-respiratory-illness.html

26. Walker K, Herman M. Tidycensus: Load US Census Boundary and Attribute Data as “Tidyverse” and “Sf”-Ready Data Frames.; 2024. https://walker-data.com/tidycensus/

27. Mistry D, Litvinova M, Pastore Y, Piontti A, et al. Inferring high-resolution human mixing patterns for disease modeling. Nat Commun. 2021;12(1):323.

28. Washington State Immunization Information System. Accessed December 13, 2024. https://doh.wa.gov/public-health-provider-resources/healthcare-professions-and-facilities/data-exchange/immunization-information-system

29. Pitzer VE, Viboud C, Alonso WJ, et al. Environmental drivers of the spatiotemporal dynamics of respiratory syncytial virus in the United States. PLoS Pathog. 2015;11(1):e1004591.

30. Hammitt Laura L., Dagan Ron, Yuan Yuan, et al. Nirsevimab for Prevention of RSV in Healthy Late-Preterm and Term Infants. N Engl J Med. 2022;386(9):837–846.

31. Walsh Edward E., Pérez Marc Gonzalo, Zareba Agnieszka M., et al. Efficacy and Safety of a Bivalent RSV Prefusion F Vaccine in Older Adults. N Engl J Med. 2023;388(16):1465–1477.

32. Papi Alberto, Ison Michael G., Langley Joanne M., et al. Respiratory Syncytial Virus Prefusion F Protein Vaccine in Older Adults. N Engl J Med. 2023;388(7):595–608.

33. Kampmann Beate, Madhi Shabir A., Munjal Iona, et al. Bivalent Prefusion F Vaccine in Pregnancy to Prevent RSV Illness in Infants. N Engl J Med. 2023;388(16):1451–1464.

34. Britton A. Use of Respiratory Syncytial Virus Vaccines in Adults Aged ≥60 Years: Updated Recommendations of the Advisory Committee on Immunization Practices — United States, 2024. MMWR Morb Mortal Wkly Rep. 2024;73. doi:10.15585/mmwr.mm7332e1

35. Dahabreh IJ, Trikalinos TA, Balk EM, Wong JB. Guidance for the Conduct and Reporting of Modeling and Simulation Studies in the Context of Health Technology Assessment. In: Methods Guide for Effectiveness and Comparative Effectiveness Reviews. Agency for Healthcare Research and Quality (US); 2016.

36. Nirsevimab Coverage, Children 0 to 19 months, United States. Accessed August 7, 2024. https://www.cdc.gov/vaccines/imz-managers/coverage/rsvvaxview/nirsevimab-coverage-children-0-19months.html

37. Perofsky AC, Hansen CL, Burstein R, et al. Impacts of human mobility on the citywide transmission dynamics of 18 respiratory viruses in pre– and post-COVID-19 pandemic years. Nat Commun. 2024;15(1):4164.

38. Falsey AR, Cameron A, Branche AR, Walsh EE. Perturbations in Respiratory Syncytial Virus Activity During the SARS-CoV-2 Pandemic. J Infect Dis. 2022;227(1):83–86.

39. Hamid S, Winn A, Parikh R, et al. Seasonality of Respiratory Syncytial Virus – United States, 2017-2023. MMWR Morb Mortal Wkly Rep. 2023;72(14):355–361.

40. Stein RT, Zar HJ. RSV through the COVID-19 pandemic: Burden, shifting epidemiology, and implications for the future. Pediatr Pulmonol. 2023;58(6):1631–1639.

41. Zheng Z, Pitzer VE, Shapiro ED, Bont LJ, Weinberger DM. Estimation of the Timing and Intensity of Reemergence of Respiratory Syncytial Virus Following the COVID-19 Pandemic in the US. JAMA Netw Open. 2021;4(12):e2141779.

42. Löwensteyn YN, Zheng Z, Rave N, et al. Year-Round Respiratory Syncytial Virus Transmission in The Netherlands Following the COVID-19 Pandemic: A Prospective Nationwide Observational and Modeling Study. J Infect Dis. 2023;228(10):1394–1399.

43. Bents SJ, Viboud C, Grenfell BT, et al. Modeling the impact of COVID-19 nonpharmaceutical interventions on respiratory syncytial virus transmission in South Africa. Influenza Other Respi Viruses. 2023;17(12):e13229.

44. Respiratory Syncytial Virus (RSV) Vaccination Coverage and Intent for Vaccination, Adults 60 Years and Older, United States. Accessed August 7, 2024. https://www.cdc.gov/vaccines/imz-managers/coverage/rsvvaxview/adults-60-coverage-intent.html

45. Respiratory Syncytial Virus (RSV) Vaccination Coverage, Pregnant Persons, United States. Accessed August 7, 2024. https://www.cdc.gov/vaccines/imz-managers/coverage/rsvvaxview/pregnant-persons-coverage-intent.html

46. Nirsevimab Receipt and Intent for Infants, United States. Accessed August 7, 2024. https://www.cdc.gov/vaccines/imz-managers/coverage/rsvvaxview/nirsevimab-coverage.html

47. Falsey AR, Hosman T, Bastian AR, et al. Long-term efficacy and immunogenicity of Ad26.RSV.preF-RSV preF protein vaccine (CYPRESS): a randomised, double-blind, placebo-controlled, phase 2b study. Lancet Infect Dis. 2024;24(9):1015–1024.

48. Vaccines for Adults Ages 60 and Over. Respiratory Syncytial Virus Infection (RSV). Accessed July 18, 2024. https://www.cdc.gov/rsv/vaccines/older-adults.html#:~:text=How%20long%20do%20these%20vaccines,for%20up%20to%202%20years.

49. Wilkins D, Yuan Y, Chang Y, et al. Durability of neutralizing RSV antibodies following nirsevimab administration and elicitation of the natural immune response to RSV infection in infants. Nat Med. 2023;29(5):1172–1179.

50. Wilson Eleanor, Goswami Jaya, Baqui Abdullah H., et al. Efficacy and Safety of an mRNA-Based RSV PreF Vaccine in Older Adults. N Engl J Med. 2023;389(24):2233–2244.

51. Oster NV, Williams EC, Unger JM, et al. Hepatitis B Birth Dose: First Shot at Timely Early Childhood Vaccination. Am J Prev Med. 2019;57(4):e117–e124.

52. Centers for Disease Control and Prevention. Evidence to Recommendations Framework (EtR): RSV Vaccination in Adults Aged 50–59 years, 60–74 years, and 75 years and older. Published online June 26, 2024. chrome-extension://efaidnbmnnnibpcajpcglclefindmkaj/https://www.cdc.gov/acip/downloads/slides-2024-06-26-28/11-RSV-Adult-Melgar-Roper-Britton-508.pdf

53. Influenza Vaccination Coverage for Persons 6 Months and Older. FluVaxView. Accessed October 15, 2024. https://www.cdc.gov/fluvaxview/interactive/general-population-coverage.html

