## Supplemental Materials for "Scenario projections of RSV hospitalizations averted due to new immunization programs in King County, Washington, October 2023 to May 2025"

Table of Contents

eMethods

eTables & eFigures

1. eFigure 1
2. eTable 1
3. eTable 2
4. eFigure 2
5. eFigure 3
6. eFigure 4
7. eFigure 5

eReferences

### eMethods

#### RSV Hospitalization Data

*Data and age groups:* We used RSV hospitalization data for King County, WA from two sources. For age groups ≥10 years we used RSV-diagnosed hospital admissions among King County residents or at King County hospitals from Washington State's syndromic surveillance platform from week 26 of 2017 through week 18 of 2024. The RSV-diagnosis codes are listed in CDC's guidance document for the National Syndromic Surveillance Program.[1] They include ICD-10, ICD-9 and SNOMED CT codes. For children <10 years we used RSV hospitalizations with a positive RSV test (during or preceding admission) seen at Seattle Children's Hospital (SCH) for the same period. We considered also using the dataset from Washington State syndromic surveillance for children<10 years but found that many admissions from SCH were missing from this data source and that there were relatively few non-SCH admissions for this age group reported in this system.

Hospitalizations were aggregated into a weekly all-ages time series and were also summarized by respiratory season (defined as week 40 of year y to week 39 of year y+1) and age group (<2m, 2-3m, 4-5m, 6-7m, 8-9m, 10-11m, 12-24m, 2-4 yrs, 5-9 yrs, 10-19 yrs, 20-39 yrs, 40-59 yrs, 60-74 yrs, ≥75 yrs). Seasonally aggregated counts allowed us to calculate the RSV age distributions for seasons before the COVID-19 pandemic (2017-2020), during the large rebound season (2022-2023), and during the most recent 2023-2024 season.

*Trends adjustments:* Reporting of RSV hospitalizations has likely changed in the wake of the COVID-19 pandemic due to increased testing for respiratory viruses. In parallel, the COVID-19 period has disrupted RSV dynamics due to social distancing, resulting in a gradual buildup of susceptibles and a large rebound season in 2022-2023. Teasing out the contribution of increased testing vs. changes in transmission intensity and resulting hospitalizations can be complicated, especially given few years of data for model calibration and potential age differences in testing. Due to these limitations, we did not fit explicit changes in reporting rates as part of our transmission modeling process. Instead, we pre-processed the weekly hospitalization data to adjust for putative changes in testing propensity over time.

We reasoned that the large rebound season in 2022-2023 could not be used for assessment of testing propensity as it was an atypical season both in timing and size, resulting from the COVID-19 immunity debt. Instead, the most recent 2023-2024 season was more typical, as exemplified by a peak timing within 5 weeks of pre-pandemic seasons. We assumed that the 2023-2024 season reflected the new RSV testing regime after the immunity debt had settled. Because RSV interventions were put in place in 2023-2024 in infants and seniors, these age groups could not be used as comparators to pre-pandemic seasons. Instead, to evaluate testing trends, we focused on those aged 1-59 years, the age groups not impacted by immunizations. We compared the average number of reported RSV hospitalizations between the pre-pandemic seasons and the 2023-2024 season, separately for the adult and pediatric groups within age brackets 1-59 yrs. Any difference between reported hospitalizations during the 2023-2024 season and pre-pandemic seasons would be due solely to changes in RSV testing and reporting propensity, rather than increased circulation or disease severity during this season. We then scaled the pre-pandemic time series and seasonal totals for all age groups by the ratios of pre-pandemic to 2023-2024 counts (the <1 yr age groups were scaled by the same value as the 1-9 yrs age group and the 60-74 years and ≥75 yrs age groups by the same value as the 10-59 yrs age group) (eFigure 1).

Our detrending approach makes the assumption that interventions in 2023-2024 did not affect the overall RSV transmission dynamics, i.e. there was no indirect effect of these interventions on unimmunized age groups. We believe this is an appropriate assumption given the modest coverage of RSV interventions in 2023-2024, the lack of evidence that RSV interventions affect infection or transmission, and the fact that the age groups targeted by RSV interventions have fairly low contact rates with the rest of the population.

**Demographic and Contact Data**

We use an open population model to simulate the population in King County, WA. We extracted age-specific population sizes, birth rates, and net migration rates for King County, WA, for the year 2022 (most recent available data) using the tidycensus R package.[2] We stratified the population into 14 age classes (<2m, 2-3m, 4-5m, 6-7m, 8-9m, 10-11m, 12-24m, 2-4 yrs, 5-9 yrs, 10-19 yrs, 20-39 yrs, 40-59 yrs, 60-74 yrs, ≥75 yrs). The annual birth rate was converted to a weekly number of births and was used to introduce new individuals into the <2m age class. The model assumes that individuals age exponentially into the next age class with the rate of aging equal to the inverse of the time spent in each age class. The net migration rate was applied uniformly across age classes. We used a contact matrix for Washington State described by Mistry et al[3] to define contacts within and between age classes.

**Immunization Data**

We obtained monthly data on RSV immunization coverage for King County, WA from July 2023 to April 2024 from Washington State's Immunization Information System (WAIIS). Data included the number of doses of nirsevimab for infants 0-7 months, the number of vaccines for adults 60-74 years and ≥75 years, and vaccine doses administered to females between 15 and 49 years. WAIIS did not have data on pregnancy status so we assumed that all doses given to females 15-49 years were maternal RSV immunizations related to pregnancy. We assumed a 1-month lag between vaccine administration during pregnancy and infant protection (i.e., doses administered to pregnant persons in September provide protection to infants born in October).

To calculate cumulative coverage proportion for maternal vaccination and birth doses of nirsevimab, we only included infants born up to a given time in the denominator (i.e., to estimate coverage proportion in November, we only included infants born in October and November in the denominator, not all infants born during the full RSV season that runs between October and March) (eTable 1). However, nirsevimab can also be given as a catch-up dose until 8 months.

For catch-up doses of nirsevimab we used a stable denominator for the cumulative coverage proportion, considering all infants <8 months at the start of the RSV immunization campaign on October 1, even though some of these infants will lose eligibility (age out) over the course of the season. Hence for catch up doses, the cumulative coverage proportion is the proportion of catch-up infants ever eligible for immunization who actually received the immunization.

We did not have data on infant age at the time of nirsevimab administration. In our main analysis we assumed that 70% of the total nirsevimab doses were administered as birth doses and 30% were administered as catch-up doses, resulting in 31.1% cumulative coverage for infants born during the RSV season and 11.4% cumulative coverage for infants eligible for a catch-up dose. This is consistent with publicly available data from Washington State where 11.4% of eligible infants born before October 1, 2023 (hence considered “catch-up”) received a dose of nirsevimab.[4] The denominator calculations and estimated coverage over time are provided in eTable 1. To use immunization coverage in our transmission model, we interpolated weekly coverage data from the monthly data.

**Immunization Effectiveness**

We used data from clinical trials and observational studies to establish model parameters related to the effectiveness of immunizations and duration of protection against hospitalization (see eTable2).

Clinical trial data for nirsevimab places the efficacy at 62.1% against RSV-related hospitalizations at 150 days post administration.[5] However, observational studies from the 2022-2023 and 2023-2024 RSV seasons have placed the effectiveness closer to 80% or higher.[6–9] Clinical trial data suggested there were differences in efficacy based on age, with lower efficacy for infants ≤3m at the time of administration (58.8% versus 92.2% for infants >3m).[5] Results from observational studies have provided mixed evidence. A study from Spain[8] found lower effectiveness among infants receiving birth doses compared to catch-up doses, while the HARMONIE[6] study found higher effectiveness among infants <3m, and a study from France[7] found no differences by age. Given the conflicting evidence, we assumed the same effectiveness for all ages, set at 80%, with a mean duration of protection of 180 days.

At the time of writing there have been no observational studies evaluating the real-world effectiveness of maternal vaccination, however, clinical trial results placed efficacy at 67.7% 90 days after birth. This declined to 56.8% 180 days after birth and was no longer significant by 360 days after birth.[10] We set the effectiveness of maternal vaccines at 57%, with a mean duration of protection of 180 days.

For both nirsevimab and maternal vaccination, we assumed that all protection was for the risk of hospitalization given infection (i.e., protection against severe disease) and that these immunizations did not prevent infection.[11]

Clinical trials for senior vaccination did not include hospitalization as an end point. The efficacy for the endpoints included ranged from 62.1 - 85.7% with more severe outcomes associated with higher efficacy estimates.[12,13] Observational data from the 2023-2024 season suggests the effectiveness against RSV-associated hospitalization was 75% for immunocompetent seniors.[14] This did not differ substantially by the vaccine used or by age group (60-74 vs. ≥ 75 yrs).[14–16] Protection from the vaccine has been shown to last for at least two years.[17] Observational data also suggested some protection against documented clinical RSV infection. In our main analysis we considered a 75% vaccine effectiveness against hospitalization lasting 730 days, and no protection against infection. In a sensitivity analysis we allowed for some protection to operate against infection and some against severe disease, assuming 50% protection against infection and 50% for hospitalization given infection.

All effectiveness parameters were treated as relative risks (RR), where:

$$RR = 1 - \frac{Vaccine\ Efficacy/Effectiveness}{100}$$

### RSV Transmission Model

We developed an age-structured compartmental RSV transmission model adapted from Pitzer et al (see **eFigure 2** for model schematic and **eTable 2** for a list of all parameters).[18] The model assumes short-term and non-sterilizing maternally-derived immunity (compartments $M_0$,$M_{n1}$,$M_{v1}$ for non-immunized infants, infants immunized by nirsevimab, and those immunized by maternal vaccination, respectively), frequent reinfections throughout life, where infectivity and recovery depends on the number of prior infections (compartments $I_1$-$I_4$), and a gradual build-up of partial immunity following infections leading to different susceptibility levels ($S_0$-$S_3$). The version of the model previously published by Pitzer et al.[18] and Zheng et al.[19] assumed no sterilizing immunity. In the adapted model described here we have accounted for a short period of sterilizing immunity by including recovered compartments following each infection ($R_1$-$R_4$). More details on this decision are provided below in the section on sensitivity analyses (eFigure 3).

In the non-immunization model, we assume that all infants are born into an "$M_0$" compartment with partial immunity against infection, with susceptibility equivalent to that of an adult who had more than 3 infections (due to the effect of maternal antibodies). After this protection wanes, infants become fully susceptible ($S_0$). Following the first infection ($I_1$) individuals have a short period of sterilizing immunity from infection ($R_1$). After this immunity wanes, individuals are susceptible again, but with a lower relative risk of infection ($S_1$). Following each infection the duration of infectiousness becomes shorter, the relative infectiousness decreases, the duration of immunity (residence in the $R_n$ compartment) increases, and the relative susceptibility decreases. The model is age specific so that all compartments are broken down by the age groups described in the previous section on demographic and contact data, with exponential aging out of each age class and a slow influx of population immigration into each class.

Interventions are modeled by separate compartments representing nirsevimab at birth ($M_{n1}$), maternal vaccine ($M_{v1}$), catch-up immunization with nirsevimab ($N_1$), and senior vaccines ($V_{s1}$).

For each of these interventions, we use an Erlang distribution that splits immunized states into two successive compartments (e.g., $M_{n1}$ and $M_{n2}$ for nirsevimab at birth) for two reasons:1) to adjust the distribution of duration of protection (with the Erlang resulting in a gamma distribution of protection, which is more realistic than exponential) and 2) to integrate the combined effects of maternal immunity and immunization products in infants (i.e., $M_{n1}$ and $M_{v1}$ compartments).

To model birth doses of nirsevimab, we assume that infants are born into compartment $M_{n1}$ at a rate consistent with birth rates in King County and nirsevimab birth uptake. Infants stay in this compartment while they enjoy the combined effects of maternal immunity and nirsevimab. After losing maternal immunity at rate $\omega_i$, infants transit to compartment $M_{n2}$, in which they are still protected by nirsevimab. Similarly, we use compartments $M_{v1}$ and $M_{v2}$ to model the outcomes of infants whose mothers were vaccinated against RSV. Catch-up infants who receive nirsevimab after birth are modeled separately, using compartments $N_1$ and $N_2$, which they enter after losing maternal immunity and at a rate reflecting catch-up immunizations in King County. To model vaccination in seniors, we use compartments $V_{s1}$ and $V_{s2}$, with individuals transiting into $V_{s1}$ based on weekly vaccine coverage in King County $\nu(t)$.

Transit out of any of these immunized compartments and into infected compartments is governed by the force of infection $\lambda_a(t)$ modulated by the degree of susceptibility corresponding to each compartment. In the main model, we assume that interventions do not affect susceptibility, so that susceptibility is only determined by the number of prior infections ($\sigma_1$-$\sigma_3$, with higher subscripts denoting reduced susceptibility after a higher number of prior infections).

The differential equations describing the transmission dynamics of RSV in the model are as follows, with *a* denoting age:

$$\frac{dM_{0,a}}{dt} = B_1(t) - \sigma_3\lambda_a(t)M_{0,a} - \omega_1 M_{0,a} - (\mu+\eta)M_{0,a}$$

$$\frac{dM_{n1,a}}{dt} = B_2(t) - \sigma_3\lambda_a(t)M_{n1,a} - \omega_i M_{n1,a} - (\mu+\eta)M_{n1,a}$$

$$\frac{dM_{n2,a}}{dt} = \omega_i M_{n1,a} - \omega_i M_{n2,a} - \lambda_a(t)M_{n2,a} - (\mu+\eta)M_{n2,a}$$

$$\frac{dM_{v1,a}}{dt} = B_3(t) - \sigma_3\lambda_a(t)M_{v1,a} - \omega_i M_{v1,a} - (\mu+\eta) M_{v1,a}$$

$$\frac{dM_{v2,a}}{dt} = \omega_i M_{v1,a} - \omega_i M_{v2,a} - \lambda_a(t)M_{v2,a} - (\mu+\eta)M_{v2,a}$$

$$\frac{dS_{0,a}}{dt} = \omega_1 M_{0,a} - \lambda_a(t)S_{0,a} - U(t)S_{0,a} - (\mu+\eta)S_{0,a}$$

$$\frac{dN_{1,a}}{dt} = \upsilon(t) - \lambda_a(t)N_{1,a} - \omega_i N_{1,a} - (\mu+\eta) N_{1,a}$$

$$\frac{dN_{2,a}}{dt} = \omega_i N_{1,a} - \lambda_a(t) N_{2,a} - \omega_i N_{2,a} - (\mu+\eta) N_{2,a}$$

$$\frac{dS_{i,a}}{dt} = \omega_i N_{2,a} + \omega_i M_{v2,a} + \omega_i M_{n2,a} - \lambda_a(t) S_{i,a} - (\mu+\eta) S_{i,a}$$

$$\frac{dI_{1,a}}{dt} = \lambda_a(t)(S_{0,a} + S_{i,a} + N_{1,a} + N_{2,a} + \sigma_3 M_{0,a} + \sigma_3 M_{n1,a} + M_{n2,a} + \sigma_3 M_{v1,a} + M_{v2,a}) - \gamma I_{1,a} - (\mu+\eta) I_{1,a}$$

$$\frac{dR_{1,a}}{dt} = \gamma I_{1,a} - \omega_2 R_{1,a} - (\mu+\eta) R_{1,a}$$

$$\frac{dS_{1,a}}{dt} = \omega_2 R_{1,a} - \sigma_1 \lambda_a(t) S_{1,a} - (\mu+\eta) S_{1,a}$$

$$\frac{dI_{2,a}}{dt} = \sigma_1 \lambda_a(t) S_{1,a} - \gamma I_{2,a} - (\mu+\eta) I_{2,a}$$

$$\frac{dR_{2,a}}{dt} = \gamma I_{2,a} - \omega_2 R_{2,a} - (\mu+\eta) R_{2,a}$$

$$\frac{dS_{2,a}}{dt} = \omega_2 R_{2,a} - \sigma_2 \lambda_a(t) S_{2,a} - (\mu+\eta) S_{2,a}$$

$$\frac{dI_{3,a}}{dt} = \sigma_2 \lambda_a(t) S_{2,a} - \gamma I_{3,a} - (\mu+\eta) I_{3,a}$$

$$\frac{dR_{3,a}}{dt} = \gamma I_{3,a} - \omega_3 R_{3,a} - (\mu+\eta) R_{3,a}$$

$$\frac{dS_{3,a}}{dt} = \omega_3 R_{3,a} + \omega_v V_{s2,a} - \sigma_3 \lambda_a(t) S_{3,a} - \nu(t) S_{3,a} - (\mu+\eta) S_{3,a}$$

$$\frac{dV_{s1,a}}{dt} = \nu(t) - \omega_v V_{s1,a} - \sigma_3 \lambda_a(t) V_{s1,a} - (\mu+\eta) V_{s1,a}$$

$$\frac{dV_{s2,a}}{dt} = \omega_v V_{s1,a} - \omega_v V_{s2,a} - \sigma_3 \lambda(t) V_{s2,a} - (\mu+\eta) V_{s2,a}$$

$$\frac{dI_{4,a}}{dt} = \sigma_3 \lambda_a(t)(S_{3,a} + V_{s1,a} + V_{s2,a}) - \gamma_3 I_{4,a} - (\mu+\eta) I_{4,a}$$

$$\frac{dR_{4,a}}{dt} = \gamma I_{4,a} - \omega_3 R_{4,a} - (\mu+\eta) R_{4,a}$$

$\lambda_a(t)$ is the age-specific force of infection on each susceptible individual in age class (a) and at time *(t)*. The force of infection varies over time following (see also ref [19]):

$$\lambda_a(t) = (1 + b1(\frac{2\pi(t-\phi)}{52.1775})) \sum_k \beta_{a,k} (I_{1,k}(t) + \rho_1 I_{2,k}(t) + \rho_2 I_{3,k}(t) + \rho_2 I_{4,k}(t) + \epsilon)/N(t)$$

Where *b1* and $\phi$ represent the amplitude and timing of the seasonal RSV peak, $\beta_{a,k}$ represents the transmission coefficient (the product of the per capita probability of transmission given contact between susceptible in age class a and infected individuals in age class *k* and the contact rate between these age groups). This is then multiplied by the number of infectious individuals of age $k$, where $\rho_1$ and $\rho_2$ denote the relative infectiousness of second and subsequent infections. Imported infections (seeding) are denoted by $\epsilon$. *N(t)* represents the population size at time *(t)*.

To calibrate the model against the data, we assume that a proportion of the incident infections (i.e., individuals moving from an S to an I compartment) require hospitalization and are recorded as RSV-hospitalizations in our data. This proportion ($\theta_{n,a}$), akin to infection severity, is influenced by both age (a) and number of previous infections (n) (see **eTable 2 for values)**.

The number of reported RSV hospitalizations at age a is given as:

$$H_a(t) = \lambda_a(t) * (\theta_{1,a}\sigma_3 M_{0,a} + \theta_{1,a}\delta_1\sigma_3 M_{n1,a} + \theta_{1,a}\delta_1 M_{n2,a} + \theta_{1,a}\delta_2\sigma_3 M_{v1,a} + \theta_{1,a}\delta_2 M_{v2,a} + \theta_{1,a} S_{0,a} + \theta_{1,a} S_{i,a} + \theta_{2,a} S_{1,a} + \theta_{3,a} S_{2,a} + \theta_{3,a} S_{3,a} + \theta_{3,a}\delta_3 V_{s1,a} + \theta_{3,a}\delta_3 V_{s2,a})$$

**Model Calibration**

We fit the model in a three-step process. In the first step, we determined the initial conditions for each model compartment. Previous RSV modeling studies have approached this by initiating the model with 1 infection in each age class >6 months and allowing the model to burn-in for 20 to 30 years before fitting to the data. We found that this approach did not preserve the age structure of our population well. Instead, we initialized the model by setting the fitted parameters to plausible values (based on results from the approach previously described), seeding one infection in each age class >6 months, and using a 200-year burn-in period to let the model reach equilibrium. For each age class, we then determined the proportion of the population in each model compartment and multiplied the age-stratified 2022 King County population by these proportions. We used these values as the initial conditions for the model. Before fitting to the data we allowed the model to burn-in for 2 additional years.

After establishing the initial conditions for each compartment, in the second step we used maximum likelihood estimation to calibrate the model to the weekly all-ages time series and age-distribution in the pre-COVID-19 period (July 2017 to April 2020). Following Pitzer et al[18], the likelihood combined a Poisson distribution for weekly hospitalizations and a multinomial distribution for the proportion of hospitalizations occurring in each age class (the 12-24 months were combined with the 2-4 years and all age groups between 5 and 59 were combined for a total of 10 age classes rather than 14). We fitted parameters for the baseline transmission rate (*q*), the amplitude of seasonal forcing (*b1*), the phase of seasonal forcing ($\phi$), and five age-specific reporting rates for the proportion of infected individuals who develop severe disease requiring (reported) hospitalization ($\theta_{n,a}$) (see eTable2). The parameters for reduced susceptibility and infectiousness, and the duration of immunity and infectiousness were fixed based on previous RSV modeling studies.[18–20]

In the final step we fit the COVID-19 pandemic period. To do this we adjusted the force of infection by fitting 3 parameters to reduce the number of contacts over periods corresponding to different interventions or behaviors (eFigure 4).[21] The timing of these periods was based on social distancing interventions in King County and reported contact patterns[22]: There was a drastic decrease in contacts during the first year of the pandemic corresponding to stay-at-home-orders, which we modeled as a reduction in contacts *c1*, relative to the pre-pandemic period; contacts resumed during the spring and fall of 2021 but did not reach normal pre-pandemic levels (modeled as contact reduction c2); contacts declined again in winter 2021-2022 due to the emergence of the omicron variant (modeled as contact reduction c3). Finally, contacts returned to pre-pandemic norms in mid-2022. Parameters c1-c3 were fitted to the data by maximizing the likelihood given all other calibrated parameters set at their estimated values from step two. We also tried fitting more granular periods of contact reductions but found that this added complexity did not result in a better model fit and led to issues with model convergence.

After calibration steps 1-3, we let the model project forward until May 2024 using the best fit parameters. These projections represent our counterfactual scenarios (no immunizations) for the 2023-2024 season. We next ran projections for the 2023-2024 season using the observed weekly coverage in King Country for maternal vaccines, infant nirsevimab, and senior vaccines, and assumptions about efficacy and duration of immunity for each of these products (see eTable 2 for assumptions). Note that in our main analyses, we assume that these immunizations reduce the risk of hospitalization given infection but do not affect risk of infection or infectiousness, and hence we do not expect indirect effects. We relax these assumptions in sensitivity analyses. We compared the results from this intervention model to our counterfactual to arrive at an estimated number of hospitalizations averted due to immunizations for the 2023-24 season. Finally, we ran projections for the 2024-2025 season under pessimistic and optimistic scenarios for immunization coverage in seniors and infants, and under a counterfactual scenario of no immunizations (see below).

To obtain projection intervals around our projections we added uncertainty to the fitted parameters. For all parameters except the seasonal phase ($\phi$) we varied point estimates by 10% from the fitted values to get upper and lower bounds around each parameter. The model was very sensitive to variation in the phase parameter $\phi$ and using a +/- 10% range on this parameter resulted in unrealistic epidemic timings. For $\phi$, we engineered the variability of the fitted parameter so that the peak of simulated trajectories for a given season fell in between that of epidemics in the pre-pandemic period and in the most recent post-pandemic season on record (for instance, to generate variability around $\phi$ for our 2023-2024 projections, we chose $\phi$ so that the peak of simulated epidemics fell between those observed in 2017-2020 and 2022-2023). This approach reflects the expectation that perturbations in RSV timing due to COVID-19 non-pharmaceutical interventions will gradually return to normal after contacts resume to baseline, and hence the return to normal will occur gradually after summer 2022. We used Latin Hypercube Sampling from the upper and lower bounds of our fitted parameters to generate 100 simulated trajectories for the model (we also tried 1000 simulated trajectories, but found there was little change in variability). We allowed for pairing between trajectories across scenarios by matching fitted parameters (i.e., trajectory 1 from the counterfactual scenario was paired with trajectory 1 from an intervention scenario, and both shared the same fitted parameters).

To assess immunization benefits, we calculate the age-specific differences in cumulative hospitalizations at the end of the season between 100 paired trajectories of intervention and counterfactual scenarios and report the median and 95% quantiles of these differences. We consider both absolute (i.e., no of hospitalizations averted per population) and relative differences (i.e., % of hospitalizations averted). For the 2023-2024 season, we used a weighting scheme to give more weight to simulations closer to observations. Weights were calculated as the inverse of the sum of the squared errors between the observed weekly all-ages RSV hospitalizations and the model projections for each intervention trajectory. This approach assumes that intervention trajectories that fall closer to data better capture the realized RSV dynamics, and hence these trajectories (along with their corresponding counterfactual) provide a more accurate estimation of immunization benefits.

**Calibrating Vaccine Effectiveness**

A previous study showed that dynamic transmission models using exponential waiting times tend to underestimate the impact of RSV vaccines compared to static cohort models.[23] The authors conclude this is likely due to conflating average efficacy (i.e., the overall efficacy reported in the clinical trial for a given period of time, generally an entire RSV season) with initial efficacy (i.e., the starting level of protection used as an input in the model, with protected individuals losing protection at an exponential rate consistent with the assumed duration of vaccine protection). Even though we used Erlang distributions with 2 waning compartments for all RSV immunizations to minimize the impact of exponentially distributed waning times, model diagnoses indicated that our realized VE was lower than expected based on the initial efficacy. To address this, we calibrated RR values that would lead to reductions in hospitalization consistent with the VE reported in clinical trials and cohort studies, following the strategy described in.[23,24] For maternal vaccination and birth doses of nirsevimab we ran a series of simulations which assumed all infants born from the first week of October through the end of March were immunized at birth, and the duration of protection was as defined in eTable 2. We tested RR values ranging from 0 (which means initial vaccine effectiveness is 100%) to 0.45 (initial effectiveness=55%) by increments of .01 and calculated the percentage reduction in hospitalizations compared to control simulations where the RR was set to 1. We then matched the simulated percentage reduction to the expected VE listed in eTable 2 and used the corresponding RR for all intervention scenarios presented in this paper. For catch-up doses of nirsevimab we used a similar approach except we assumed all eligible individuals received their immunization the first week of October. We used the same approach for older adult vaccines but found that there was no discrepancy between the realized and expected VE. This is likely due to our assumption of a 2-year duration of protection, which is substantially longer than the RSV season (so that exit out of the vaccine-protected state during the RSV season will be negligible). For older adults, we did adjust the vaccine coverage to account for the particular structure of our model, where only susceptible individuals are vaccinated. In reality, some individuals infected in the prior season and in the recovered state (R compartment, and specifically R4 for older individuals) will be vaccinated. Vaccine doses given to individuals in the R compartment will be “wasted” since we assume sterilizing immunity in this compartment. To account for this, we reduced the number of administered vaccines in the S3 compartment by the proportion of individuals in the R4 compartment at the start of the season for each age group, 60-74 years and ≥75 years, respectively. See eTable 2 and eFigure 5 for initial and average parameter values.

**Scenarios for the 2024-25 Season**

Following the approach taken by the RSV Scenario Modeling Hub during the 2023-2024 season (https://github.com/midas-network/covid19-scenario-modeling-hub) we established optimistic and pessimistic scenarios for immunization coverage in seniors and infants during the 2024-2025 season, representing four scenarios plus a counterfactual non-intervention scenario. In the counterfactual scenario we assumed that there was no immunization of either infants or older adults in either 2023-2024 or 2024-2025. In the pessimistic scenarios we assumed that the total coverage in adults ≥60 years was 37.5% (which includes those vaccinated in 2023-2024, since vaccine protection was expected to last over 2 RSV seasons), and that the coverage in infants was 40% for maternal vaccination and nirsevimab combined (see Table 1 in main text). In the optimistic scenarios we assumed that the total coverage in adults ≥60 years was 50%. For infants, we assumed combined coverage reached 80%, based on birth dose coverage of the hepatitis B vaccine in Washington State.[25]

In sensitivity analysis, we examined alternative strategies for infant immunization, using the most optimistic scenario - Scenario A - as reference. In scenario A2 the total infant coverage remained at 80%, but we extended the duration of protection for nirsevimab from 180 days to 270 days. In scenario A3, all catch-up doses of nirsevimab were administered from October to November 2024, rather than in a slower ramp up throughout the season. In scenario A4 all protection was given via nirsevimab (no maternal vaccination). In scenario A5, we assumed all newborns were protected by maternal vaccination (no nirsevimab birth doses).

We also ran sensitivity analyses for adults ≥60 years. In our main analysis, we assumed that the vaccine remained effective for 2 years and that vaccine protection was against hospitalization given infection but did not prevent infection. In sensitivity analysis we considered scenarios where the protection in older adults was reduced by 50% in the second year after administration (i.e., VE in the second year=75%*0.5=37.5%). We also ran analyses where protection was split equally between protection against infection and protection against hospitalization given infection (resulting in the same overall VE of 75%).

**Sensitivity Analyses - Immunity Structures**

We ran a number of sensitivity analyses testing the immunity structure of the model. The initial model we adapted from Pitzer et al[18] did not include any sterilizing immunity (no R compartment) following infections. Initial results indicated that including R compartments was important to reproduce the post COVID-19 pandemic dynamics of RSV, particularly during the large rebound seasons. To evaluate the importance of the R compartment, we fit the initial version of the model published by Pitzer et al[18] (referred to as MSIS model) and a similar model which included R compartments following each infection (referred to as MSIRS model). We fixed the infant reporting rates to be similar in both models (using the values from Zheng et al[19]) and then fit the remaining model parameters using MLE. Using the best-fitted parameters we reduced contacts by 40% from April 1, 2020 - April 1, 2021, to suppress RSV transmission during the 2020-2021 winter season and then restored contacts to typical pre-pandemic levels. Results are shown in (eFigure 3a). We found that the MSIRS model resulted in a large RSV epidemic immediately after contact rates returned to normal. The MSIS model, however, did not have an RSV epidemic until the typical timing of the next winter season. This suggests that population level immunity and a build-up of susceptibles is partially driving RSV epidemics in the MSIRS model while the MSIS model is driven more by seasonal forcing. Because the MSIRS model more accurately reproduced the post-pandemic rebound, especially out-of-season RSV activity, and timing of RSV activity is important to capture as it interacts with the timing of immunization, we used the MSIRS model in our final analysis. We note that this is not the only approach for building up a pool of susceptibles during the pandemic. Authors from the Netherlands noted similar difficulties in recreating the pandemic and rebound patterns using an MSIS model and allowed for waning into earlier S compartments (S3 wanes to S2, S2 to S1, etc.).[26] We chose to include an R compartment because it is a common feature of other RSV models.[20,21,27] In reality much remains unknown about RSV immunity, and it is difficult to know which model structure more accurately reflects the true dynamics of immunity.

In addition to the time-limited sterilizing immunity provided by the R compartments, the model also provides for sustained reductions in susceptibility. Using the version of the model with R compartments, we tested the reduced susceptibility parameters used in the Pitzer model versus those used by Hodgson et al.[20] The Hodgson et al. parameters preserve more susceptibility following the first and second infections with a larger drop between the second and third infections compared to Pitzer et al. We found that the Hodgson parameters resulted in a steeper drop off in attack rates between children, particularly those <2 years of age, compared to adults (eFigure 3b and 3c). While RSV infection data is limited, this decreasing attack rate with age is more in line with our expectations based on household and cohort studies.[28,29] Hence we used the Hodgson et al. parameters in our final model.

**eFigure1. Time series of RSV-coded hospitalizations in King County, WA.** To adjust for changes in RSV testing and reporting we compared the average number of RSV hospitalizations among those aged 1-59 years of age (the age groups not impacted by immunizations) during the 2023-2024 season with hospitalizations reported during the pre-pandemic seasons. We then scaled the pre-pandemic time series and seasonal totals up by these amounts, using different scaling for individuals < and ≥ 10 yrs. The gray area represents raw data. The orange line represents the data after rescaling and the navy line represents the rescaled data with a 3-week moving average. We used this smooth data to fit the model.

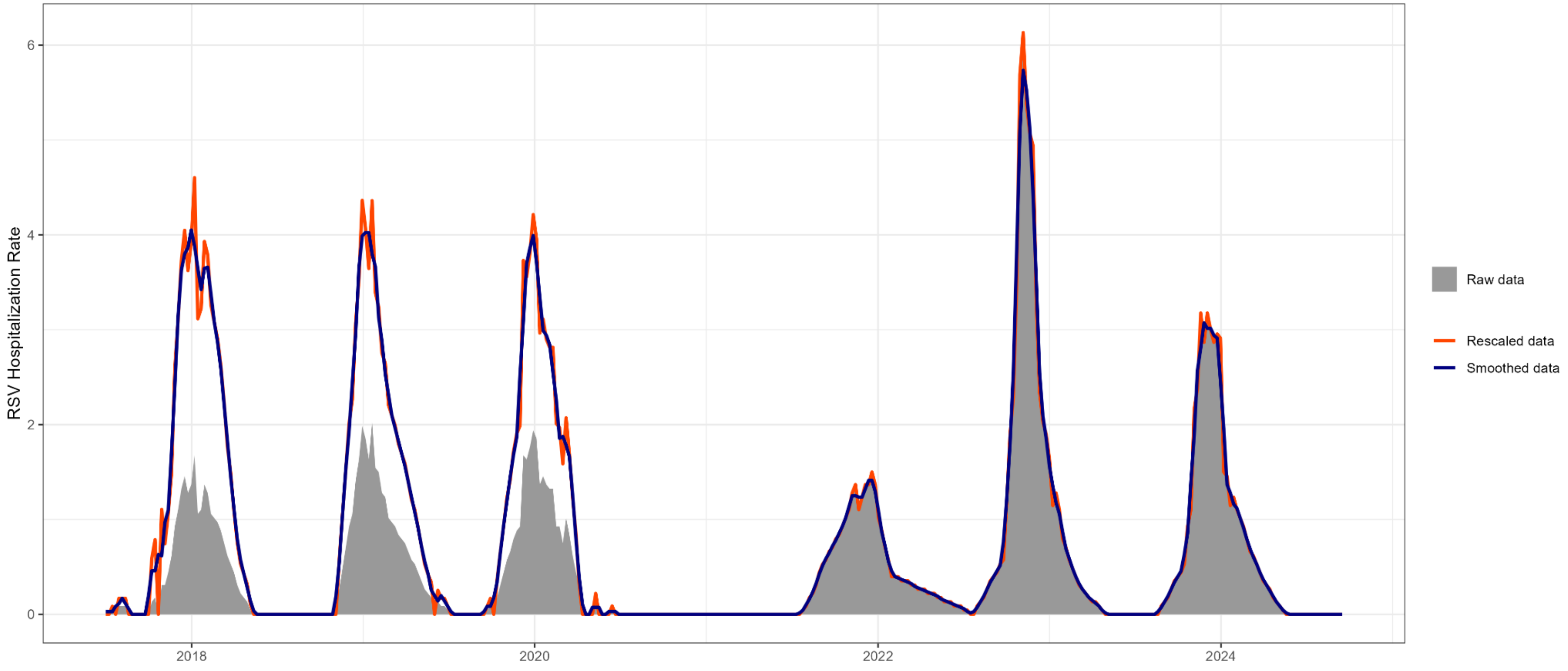

**eTable 1. Reported Infant immunizations in 2023-24 and population denominators.** Green cells represent infants born during the RSV season (using the average number of monthly births in 2022). These infants are included in the denominator calculation for both maternal vaccination and birth doses of nirsevimab. Blue cells represent the infants <8 months who were born prior to October 1, 2023, and are eligible to receive a catch-up dose of nirsevimab. Gray cells represent the infants who have aged out of nirsevimab eligibility, but were eligible at one point during the season and are included in the denominator of infants ever eligible for nirsevimab catch-up doses. We assume a 1-month delay between maternal vaccine administration and infant protection.

| **Birth month (age in months)** | **October** | **November** | **December** | **January** | **February** | **March** |
|---|---|---|---|---|---|---|
| October (0) | 1980.2 | 1980.2 | 1980.2 | 1980.2 | 1980.2 | 1980.2 |
| September (1) | 1980.2 | 1980.2 | 1980.2 | 1980.2 | 1980.2 | 1980.2 |
| August (2) | 1980.2 | 1980.2 | 1980.2 | 1980.2 | 1980.2 | 1980.2 |
| July (3) | 1980.2 | 1980.2 | 1980.2 | 1980.2 | 1980.2 | 1980.2 |
| June (4) | 1980.2 | 1980.2 | 1980.2 | 1980.2 | 1980.2 | 1980.2 |
| May (5) | 1980.2 | 1980.2 | 1980.2 | 1980.2 | 1980.2 | 1980.2 |
| April (6) | 1980.2 | 1980.2 | 1980.2 | 1980.2 | 1980.2 | 1980.2 |
| March (7) | 1980.2 | 1980.2 | 1980.2 | 1980.2 | 1980.2 | 1980.2 |
| March (8) | | 1980.2 | 1980.2 | 1980.2 | 1980.2 | 1980.2 |
| March (9) | | | 1980.2 | 1980.2 | 1980.2 | 1980.2 |
| March (10) | | | | 1980.2 | 1980.2 | 1980.2 |
| March (11) | | | | | 1980.2 | 1980.2 |
| March (12) | | | | | | 1980.2 |
| Cumulative infants born October - March | 1980.2 | 3960.4 | 5940.6 | 7920.8 | 9901 | 11881.2 |
| Cumulative infants born March - October (eligible for catch-up dose) | 13861.4 | 13861.4 | 13861.4 | 13861.4 | 13861.4 | 13861.4 |

| | | | | | | |
|---|---|---|---|---|---|---|
| Cumulative infants eligible for immunization | 15841.6 | 17821.8 | 19802 | 21782.2 | 23762.4 | 25742.6 |
| **Maternal Vaccination** | | | | | | |
| Cumulative infants born to vaccinated persons | 17 | 385 | 1098 | 2004 | 2933 | 3214 |
| Cumulative % Protected by maternal vaccination | 0.9 | 9.7 | 18.5 | 25.3 | 29.6 | 27.1 |
| **Nirsevimab Birth Doses** | | | | | | |
| Nirsevimab birth doses (70% of total doses) | 827 | 1700 | 2243 | 2685 | 3310 | 3699 |
| Cumulative % protected by nirsevimab birth dose | 41.8 | 42.9 | 37.8 | 33.9 | 33.4 | 31.1 |
| **Nirsevimab Catch-up Doses** | | | | | | |
| Nirsevimab catch-up doses (30% of total doses) | 354 | 729 | 961 | 1151 | 1419 | 1585 |
| Cumulative % of infants ever protected by nirsevimab catch-up dose | 2.6 | 5.3 | 6.9 | 8.3 | 10.2 | 11.4 |
| **Nirsevimab Total** | | | | | | |
| Total Nirsevimab doses (as reported by IIS) | 1181 | 2429 | 3204 | 3836 | 4729 | 5284 |
| Cumulative % of infants ever protected by nirsevimab | 7.5 | 13.6 | 16.2 | 17.6 | 19.9 | 20.5 |
| **Any Method** | | | | | | |
| Cumulative infants eligible for any immunization | 15841.6 | 17821.8 | 19802 | 21782.2 | 23762.4 | 25742.6 |
| Cumulative doses of any immunization | 1198 | 2814 | 4302 | 5840 | 7662 | 8498 |
| Cumulative % protected by any method | 7.6 | 15.8 | 21.7 | 26.8 | 32.2 | 33.0 |

**eTable 2. Model Parameters.** Some parameters are fixed based on the literature and others are calibrated, as indicated in the last column. We fitted parameters for the baseline transmission rate, the amplitude of seasonal forcing, the phase of seasonal forcing, and five age-specific reporting rates for the proportion of infected individuals who develop severe disease requiring (reported) hospitalization. To fit the COVID-19 pandemic period we adjusted the force of infection to account for suppressed RSV transmission by fitting 3 parameters to reduce the number of contacts, as shown in eFigure 4.

| Description | Symbol | Value | Ref |
|---|---|---|---|
| Transmission parameter | $q$ | 8.54 (7.69 - 9.40) | Fitted |
| Amplitude of seasonal forcing | $b_1$ | 0.21 (0.18 - 0.23) | Fitted |
| Phase of seasonal forcing (2023-2024 Season) | $\phi$ | 5.53 (5.34 - 5.53) | Fitted |
| Phase of seasonal forcing (2023-2024 Season) | $\phi$ | 5.53 (5.39 - 5.53) | Fitted |
| Weekly number of seeded infections | $\epsilon$ | 22.67 | 1 per 100K population |
| Relative risk of infection following first infection | $\sigma_1$ | 0.89 | 20 |
| Relative risk of infection following second infection | $\sigma_2$ | 0.72 | 20 |
| Relative risk of infection following third and subsequent infections | $\sigma_3$ | 0.24 | 20 |
| Duration of Infectiousness (days) during first infection | $1/\gamma_1$ | 10 | 18,19 |
| Duration of Infectiousness (days) during second infection | $1/\gamma_2$ | 7 | 18,19 |
| Duration of Infectiousness (days) during third and subsequent infections | $1/\gamma_3$ | 5 | 18,19 |
| Relative infectiousness of second infection (compared to naive) | $\rho_1$ | 0.75 | 18,19 |
| Relative infectiousness third and subsequent infections | $\rho_2$ | 0.51 | 18,19 |
| Duration of trans-placentally derived immunity (days) | $1/\omega_1$ | 90 | 18–20,30 |
| Relative risk of infection while protected by trans-placentally derived immunity | $\sigma_3$ | 0.24 | 20 (assuming same level of protection as S3 compartment) |

| | | | |
|---|---|---|---|
| Duration of immunity following first and second infections (days) | $1/\omega_2$ | 182.625 | [31] |
| Duration of immunity following third and subsequent infections (days) | $1/\omega_3$ | 358.9 | [20] |
| Proportion of contacts relative to the pre-pandemic period, April 2020 - April 2021 | c1 | 0.39 (0.35 - 0.43) | Fitted |
| Proportion of contacts relative to the pre-pandemic period, September 2021 - November 2021 | c2 | 0.80 (0.72 - 0.88) | Fitted |
| Proportion of contacts relative to the pre-pandemic period, December 2021 - March 2022 | c3 | 0.53 (0.47 - 0.58) | Fitted |
| Proportion of infections leading to reported hospitalization (first infection) | | | |
| <2m | $\theta_1$ | 0.085 (0.077 - 0.094) | Fitted |
| 2-3m | $\theta_1$ | 0.050 (0.045 - 0.055) | 0.085* 0.59 [19] |
| 4-5m | $\theta_1$ | 0.028 (0.025 - 0.031) | 0.085* 0.33 [19] |
| 6-7m | $\theta_1$ | 0.017 (0.015 - 0.019) | 0.085* 0.20 [19] |
| 8-9m | $\theta_1$ | 0.013 (0.012 - 0.014) | 0.085* 0.15 [19] |
| 10-11m | $\theta_1$ | 0.013 (0.012 - 0.014) | 0.085* 0.15 [19] |
| 1-4yrs | $\theta_1$ | 0.018 (0.016 - 0.020) | Fitted |
| ≥5yrs | $\theta_1$ | 0.001 | [19] |
| Proportion of infections leading to reported hospitalizations (second infection) | $\theta_2$ | $\theta_1$*0.4 | [19] |
| Proportion of infections leading to reported hospitalizations (third infection) | | | |

| | | | |
|---|---|---|---|
| <5 yrs | $\theta_3$ | 0.001 | 19 (assuming similar to first infection in ≥5 yrs) |
| 5-59 yrs | $\theta_3$ | 0.0004 (0.0004-0.0005) | Fitted |
| 6-74 yrs | $\theta_3$ | 0.003 (0.003 - 0.003) | Fitted |
| ≥75 yrs | $\theta_3$ | 0.018 (0.016 - 0.020) | Fitted |
| Immunization Parameters | | | |
| Average number of weekly births in King County, WA | Births | 455.4 | 2 |
| Birth rate for newborns who are not protected by immunizations at birth | $B_1$ | Scenario based | |
| Birth rate for newborns who receive monoclonal antibodies at birth | $B_2$ | Scenario based | |
| Birth rate for newborns who are born to vaccinated persons | $B_3$ | Scenario based | |
| Immunization rate for infants receiving catch-up doses of monoclonal antibodies. | $u$ | Scenario based | |
| Average relative risk of hospitalization for infants receiving monoclonal antibodies [VE=(1-RR)*100] (see eMethods, eFigure 5) | $\delta_1$ | 0.2 | 6,7,32 |
| Average relative risk of hospitalization for infants born to vaccinated mothers [VE=(1-RR)*100] (see eMethods, eFigure 5) | $\delta_2$ | 0.43 | 10,33 |
| Duration of protection from monoclonal antibodies and vaccination in pregnant women (days) | $2*(1/\omega_i)$ | 180 | 10,33 |
| Vaccination rate for adults ≥60 years. | $\nu$ | Scenario based | |
| Average relative risk of hospitalization for vaccinated adults ≥60 years [VE=(1-RR)*100] (see eMethods) | $\delta_3$ | 0.25 | 12–16,34 |
| Duration of protection from vaccination in adults ≥60 years | $2*(1/\omega_v)$ | 730.5 | 16,17 |

**eFigure 2. Model Diagram.** The model assumes short term maternally-derived immunity (compartments $M_0$, $Mn_1$,$M_{v1}$), frequent reinfection throughout life (compartments $I_1$-$I_4$), a short period of sterilizing immunity following infection ($R_1$-$R_4$), and a gradual build-up of partial immunity following infections ($S_0$-$S_3$). All infants are born into an "M" compartment with partial immunity against infection for 90 days. Infants born to vaccinated mothers ($M_{v1}$) or receiving a birth dose of nirsevimab ($M_{n1}$) have this protection against infection and additional protection against hospitalization given infection. When the protection against infection wanes, they continue to have protection against hospitalization for another 90 days ($M_{v2}$, $M_{n2}$). After this protection wanes, infants become fully susceptible ($S_0$). Infants receiving a catch-up dose of nirsevimab ($N_1$, $N_2$) do not have any protection against infection but do have protection against hospitalization given infection. Following the first infection ($I_1$) individuals have a short period of immunity from infection ($R_1$). After this immunity wanes, individuals are susceptible again, but with a lower relative risk of infection. Following each infection, the duration of infectiousness becomes shorter, the relative infectiousness decreases, the duration of immunity increases, and the relative susceptibility is lower. In the main analyses older adults who receive a vaccine ($V_{s1}$, $V_{s2}$) do not have any protection against infection only protection against hospitalization given infection.

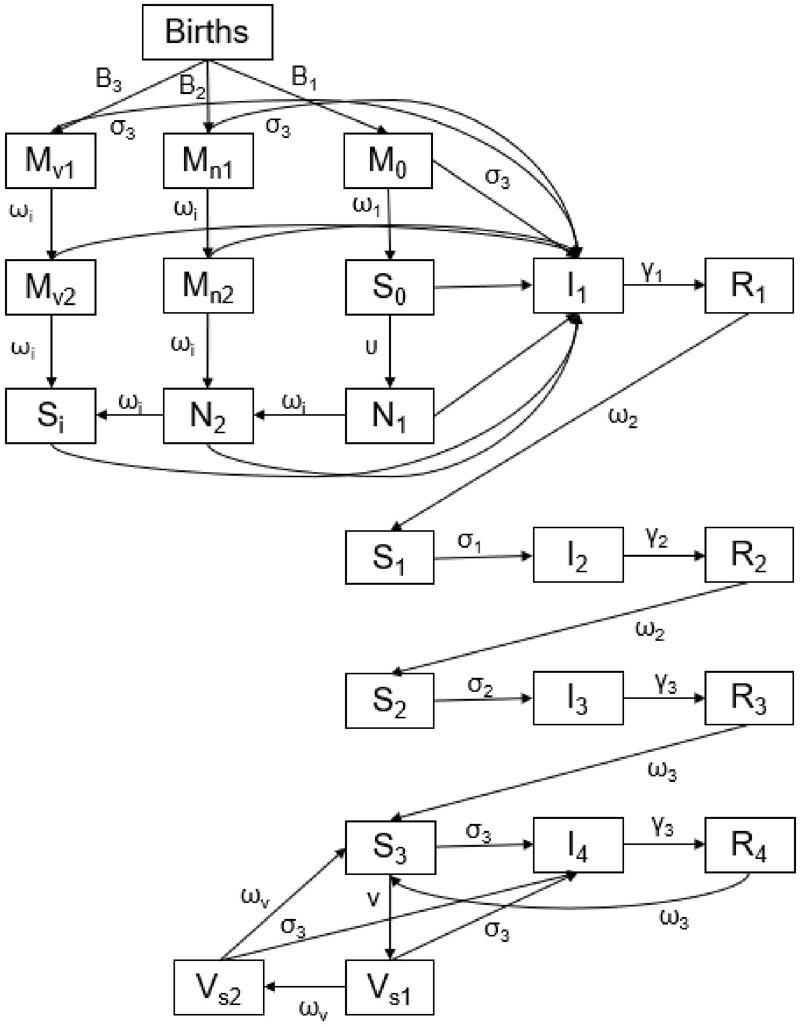

**eFigure 3. Sensitivity Analysis - Immunity Structures.** Panel A) shows our comparison between the MSIS and MSIRS models. Using the best-fitted parameters we reduced contacts by 40% from April 1, 2020 - April 1, 2021, to suppress RSV transmission during the 2020-2021 winter season and then restored them to typical pre-pandemic levels (black line). We found that the MSIRS model (orange line) resulted in an RSV epidemic immediately after contact rates returned to normal. The MSIS model (blue line), however, did not have an RSV epidemic until the typical timing of the winter season. Panel B) shows the attack rates when using the reduced susceptibility parameters from Pitzer et al. Panel C) shows the attack rates using the reduced susceptibility parameters from Hodgson et al.

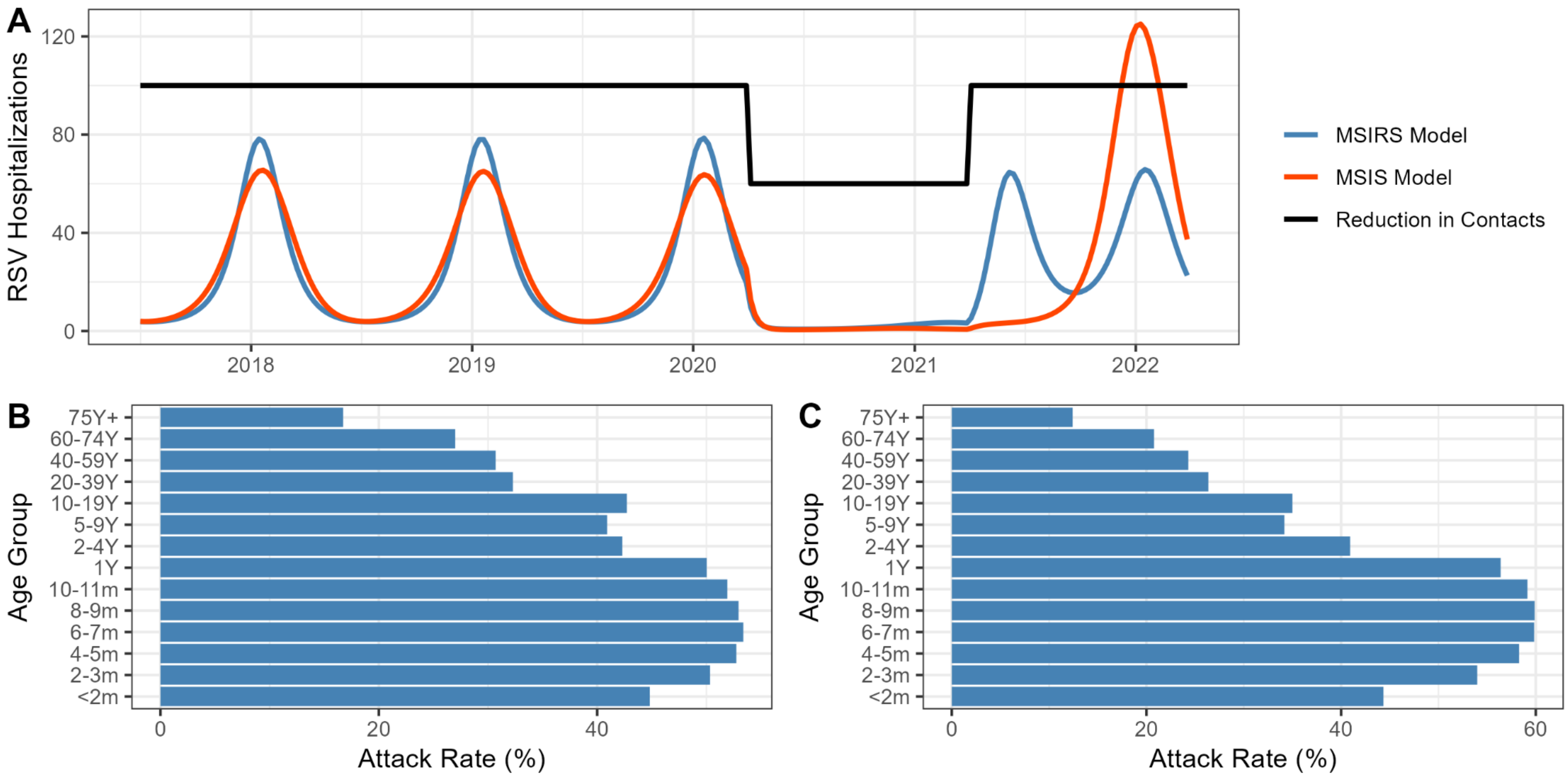

**eFigure 4. Calibration of RSV hospitalizations during the pandemic and post-pandemic periods** We calibrated the model using data from July 2017 to September 2023. The gray circles show the rescaled weekly hospitalization rates we used for calibration. The blue line shows the model fit to the data (left y-axis). The solid black line shows the estimated reduction in contacts for 3 periods of the COVID-19 pandemic. Contact reductions are relative to pre-pandemic baseline contacts (right y-axis).

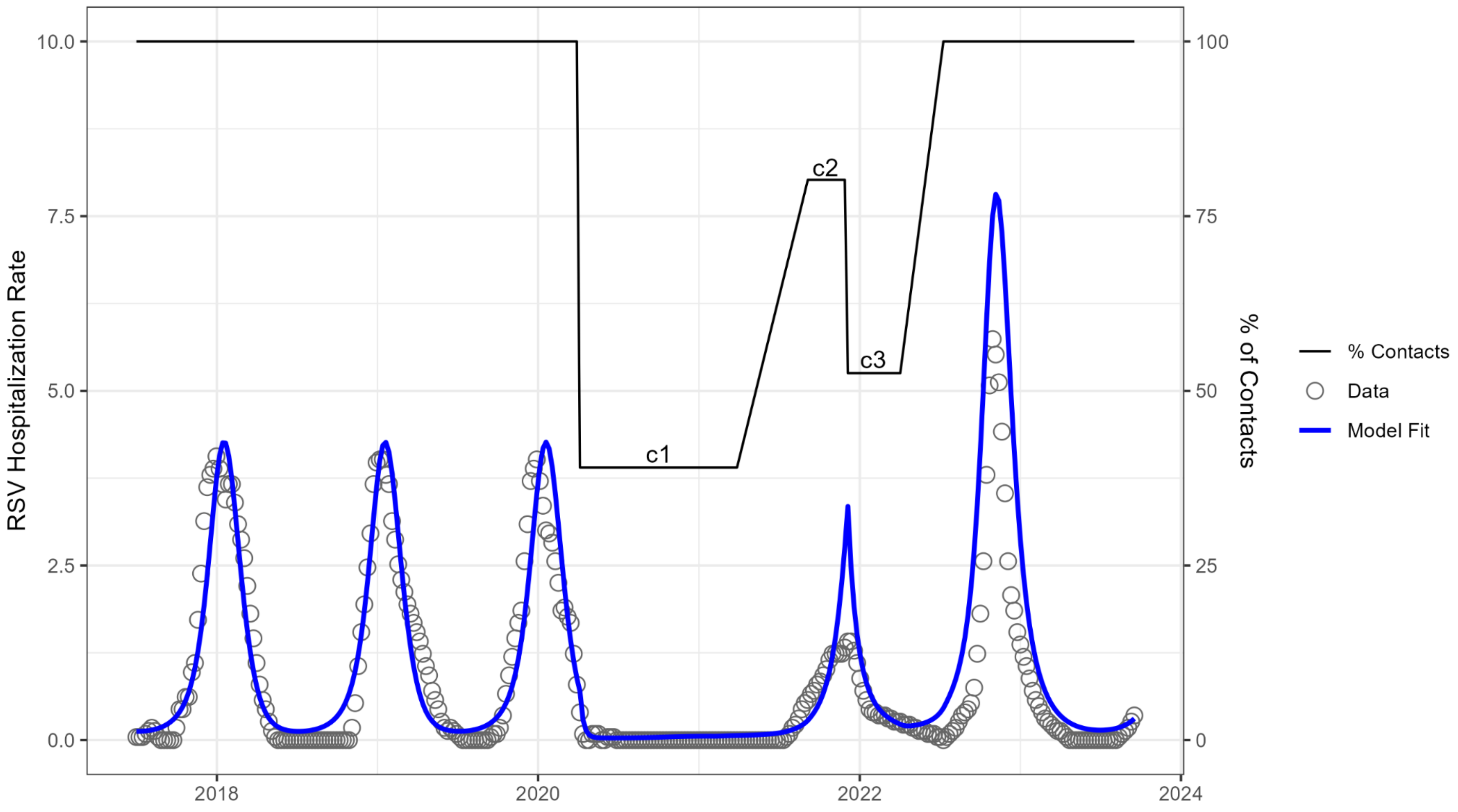

**eFigure5. Starting values for immunization effectiveness.** We identified starting values for immunization effectiveness that would lead to reductions in hospitalization consistent with clinical trials and cohort studies: 80% over 180 days for nirsevimab and 57% over 180 days for maternal vaccination. Panel A) For maternal vaccination and birth doses of nirsevimab we ran a series of simulations which assumed all infants born from the first week of October through the end of March were immunized. We tested RR values ranging from 0 (which means initial vaccine effectiveness is 100%) to 0.45 (initial effectiveness=55%) by increments of .01 and calculated the percentage reduction in hospitalizations compared to a control group where the RR was set to 1. Panel B) For catch-up doses of nirsevimab we used a similar approach, except we assumed all eligible infants received their immunization the first week of October. We selected the RR values that resulted in hospitalization reductions consistent with those observed in trials. For nirsevimab we found that an initial value of 0.01 (which translates to 99% effectiveness) resulted in a roughly 80% reduction (dark blue line) for both birth doses (82%) and catch-up doses (78%). For maternal vaccination an initial value of 0.3 (which translates to 70% effectiveness) resulted in a 57% reduction (orange line).

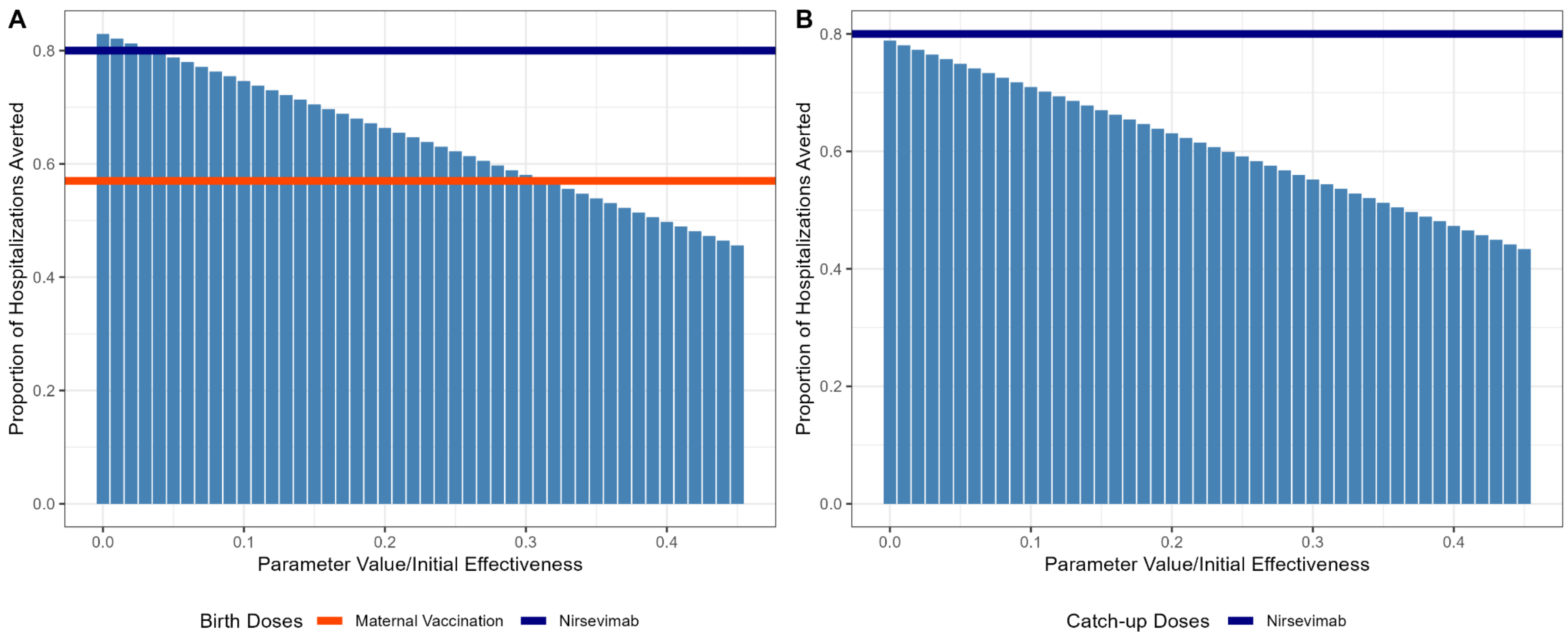

**eREFERENCES**

1. CDC. Companion Guide: NSSP ED Data on Respiratory Illness. Accessed October 4, 2024. https://www.cdc.gov/nssp/php/onboarding-resources/companion-guide-ed-data-respiratory-illness.html

2. Walker K, Herman M. *Tidycensus: Load US Census Boundary and Attribute Data as "Tidyverse" and "Sf"-Ready Data Frames.*; 2024. https://walker-data.com/tidycensus/

3. Mistry D, Litvinova M, Pastore Y Piontti A, et al. Inferring high-resolution human mixing patterns for disease modeling. *Nat Commun*. 2021;12(1):323.

4. Nirsevimab Coverage, Children 0 to 19 months, United States. Accessed August 7, 2024. https://www.cdc.gov/vaccines/imz-managers/coverage/rsvvaxview/nirsevimab-coverage-children-0-19months.html

5. Hammitt Laura L., Dagan Ron, Yuan Yuan, et al. Nirsevimab for Prevention of RSV in Healthy Late-Preterm and Term Infants. *N Engl J Med*. 2022;386(9):837-846.

6. Drysdale Simon B., Cathie Katrina, Flamein Florence, et al. Nirsevimab for Prevention of Hospitalizations Due to RSV in Infants. *N Engl J Med*. 2023;389(26):2425-2435.

7. Assad Zein, Romain Anne-Sophie, Aupiais Camille, et al. Nirsevimab and Hospitalization for RSV Bronchiolitis. *N Engl J Med*. 2024;391(2):144-154.

8. Ares-Gómez S, Mallah N, Santiago-Pérez MI, et al. Effectiveness and impact of universal prophylaxis with nirsevimab in infants against hospitalisation for respiratory syncytial virus in Galicia, Spain: initial results of a population-based longitudinal study. *Lancet Infect Dis*. Published online April 30, 2024. doi:10.1016/S1473-3099(24)00215-9

9. Moline HL, Tannis A, Toepfer AP, et al. Early Estimate of Nirsevimab Effectiveness for Prevention of Respiratory Syncytial Virus-Associated Hospitalization Among Infants Entering Their First Respiratory Syncytial Virus Season - New Vaccine Surveillance Network, October 2023-February 2024. *MMWR Morb Mortal Wkly Rep*. 2024;73(9):209-214.

10. Kampmann Beate, Madhi Shabir A., Munjal Iona, et al. Bivalent Prefusion F Vaccine in Pregnancy to Prevent RSV Illness in Infants. *N Engl J Med*. 2023;388(16):1451-1464.

11. Wilkins D, Yuan Y, Chang Y, et al. Durability of neutralizing RSV antibodies following nirsevimab administration and elicitation of the natural immune response to RSV infection in infants. *Nat Med*. 2023;29(5):1172-1179.

12. Walsh Edward E., Pérez Marc Gonzalo, Zareba Agnieszka M., et al. Efficacy and Safety of a Bivalent RSV Prefusion F Vaccine in Older Adults. *N Engl J Med*. 2023;388(16):1465-1477.

13. Papi Alberto, Ison Michael G., Langley Joanne M., et al. Respiratory Syncytial Virus Prefusion F Protein Vaccine in Older Adults. *N Engl J Med*. 2023;388(7):595-608.

14. Surie D, Self WH, Zhu Y, et al. RSV vaccine effectiveness against hospitalization among

US adults 60 years and older. *JAMA*. Published online September 4, 2024. doi:10.1001/jama.2024.15775

15. Surie D. Effectiveness of adult respiratory syncytial virus (RSV) vaccines, 2023–2024. Presented at: ACIP Presentation Slides: June 26-28, 2024 Meeting; June 26, 2024. https://www.cdc.gov/vaccines/acip/meetings/downloads/slides-2024-06-26-28/07-RSV-Adult-Surie-508.pdf

16. Britton A. Use of Respiratory Syncytial Virus Vaccines in Adults Aged ≥60 Years: Updated Recommendations of the Advisory Committee on Immunization Practices — United States, 2024. *MMWR Morb Mortal Wkly Rep*. 2024;73. doi:10.15585/mmwr.mm7332e1

17. Vaccines for Adults Ages 60 and Over. Respiratory Syncytial Virus Infection (RSV). Accessed July 18, 2024. https://www.cdc.gov/rsv/vaccines/older-adults.html#:~:text=How%20long%20do%20these%20vaccines,for%20up%20to%202%20years.

18. Pitzer VE, Viboud C, Alonso WJ, et al. Environmental drivers of the spatiotemporal dynamics of respiratory syncytial virus in the United States. *PLoS Pathog*. 2015;11(1):e1004591.

19. Zheng Z, Weinberger DM, Pitzer VE. Predicted effectiveness of vaccines and extended half-life monoclonal antibodies against RSV hospitalizations in children. *NPJ Vaccines*. 2022;7(1):127.

20. Hodgson D, Pebody R, Panovska-Griffiths J, Baguelin M, Atkins KE. Evaluating the next generation of RSV intervention strategies: a mathematical modelling study and cost-effectiveness analysis. *BMC Med*. 2020;18(1):348.

21. Bents SJ, Viboud C, Grenfell BT, et al. Modeling the impact of COVID-19 nonpharmaceutical interventions on respiratory syncytial virus transmission in South Africa. *Influenza Other Respi Viruses*. 2023;17(12):e13229.

22. Perofsky AC, Hansen CL, Burstein R, et al. Impacts of human mobility on the citywide transmission dynamics of 18 respiratory viruses in pre- and post-COVID-19 pandemic years. *Nat Commun*. 2024;15(1):4164.

23. Lang JC, Kura K, Garba SM, Elbasha EH, Chen YH. Comparison of a static cohort model and dynamic transmission model for respiratory syncytial virus intervention programs for infants in England and Wales. *Vaccine*. 2024;42(8):1918-1927.

24. Hodgson D, Wilkins N, van Leeuwen E, et al. Protecting infants against RSV disease: an impact and cost-effectiveness comparison of long-acting monoclonal antibodies and maternal vaccination. *Lancet Reg Health Eur*. 2024;38(100829):100829.

25. Oster NV, Williams EC, Unger JM, et al. Hepatitis B Birth Dose: First Shot at Timely Early Childhood Vaccination. *Am J Prev Med*. 2019;57(4):e117-e124.

26. Löwensteyn YN, Zheng Z, Rave N, et al. Year-Round Respiratory Syncytial Virus Transmission in The Netherlands Following the COVID-19 Pandemic: A Prospective Nationwide Observational and Modeling Study. *J Infect Dis*. 2023;228(10):1394-1399.

27. Hogan AB, Campbell PT, Blyth CC, et al. Potential impact of a maternal vaccine for RSV: A mathematical modelling study. *Vaccine*. 2017;35(45):6172-6179.

28. Cooney MK, Fox JP, Hall CE. The Seattle Virus Watch. VI. Observations of infections with and illness due to parainfluenza, mumps and respiratory syncytial viruses and Mycoplasma pneumoniae. *Am J Epidemiol*. 1975;101(6):532-551.

29. Cohen C, Kleynhans J, Moyes J, et al. Incidence and transmission of respiratory syncytial virus in urban and rural South Africa, 2017-2018. *Nat Commun*. 2024;15(1):116.

30. Chu HY, Steinhoff MC, Magaret A, et al. Respiratory syncytial virus transplacental antibody transfer and kinetics in mother-infant pairs in Bangladesh. *J Infect Dis*. 2014;210(10):1582-1589.

31. Ohuma EO, Okiro EA, Ochola R, et al. The natural history of respiratory syncytial virus in a birth cohort: the influence of age and previous infection on reinfection and disease. *Am J Epidemiol*. 2012;176(9):794-802.

32. Jones JM, Fleming-Dutra KE, Prill MM, et al. Use of Nirsevimab for the Prevention of Respiratory Syncytial Virus Disease Among Infants and Young Children: Recommendations of the Advisory Committee on Immunization Practices - United States, 2023. *MMWR Morb Mortal Wkly Rep*. 2023;72(34):920-925.

33. Fleming-Dutra KE, Jones JM, Roper LE, et al. Use of the Pfizer Respiratory Syncytial Virus Vaccine During Pregnancy for the Prevention of Respiratory Syncytial Virus-Associated Lower Respiratory Tract Disease in Infants: Recommendations of the Advisory Committee on Immunization Practices - United States, 2023. *MMWR Morb Mortal Wkly Rep*. 2023;72(41):1115-1122.

34. National Center for Immunization and Respiratory Diseases. Healthcare Providers: RSV Vaccination for Adults 60 Years of Age and Over. July 3, 2024. Accessed July 18, 2024. https://www.cdc.gov/vaccines/vpd/rsv/hcp/older-adults.html#:~:text=AREXVY%20was%20approximately%2077%25%20to,in%20adults%2060%20and%20older.
